# Uncovering research patterns of a middle-income country: A Comprehensive Study of Peru’s Specialized Health Institutes’ Scientific Output (1991-2021)

**DOI:** 10.1101/2025.03.03.25323252

**Authors:** Fernanda Barriga-Chambi, Frank Zela-Coila, Judith Marie Merma-Valero, Edison Salvador-Oscco, Glenny Alexandra Mendoza-Anco, Beatriz M. Luque-Mamani, Edwin Garcia-Vasquez, Carlos Quispe-Vicuña, Cender U. Quispe-Juli

## Abstract

**Objetive:**This study aimed to describe the scientific output of specialized health institutes in Peru from 1991 to 2021. **Method:** A bibliometric analysis was conducted to evaluate the scientific production, collaboration, and impact of nine specialized health institutes in Peru. An exploratory analysis of associations was performed for highly cited articles and publications in Q1 journals. Results: A total of 3,020 articles were analyzed, with the majority published in English (61.39%). The institutes with the highest scientific output were the Instituto Nacional de Salud, Instituto Nacional de Enfermedades Neoplásicas, and Instituto Nacional de Salud del Niño - Breña. Instituto Nacional de Salud collaborated with the other institutes at least once, while Instituto Nacional de Oftalmología had the least collaboration. The three institutes with the highest bibliometric impact were the Instituto Nacional de Salud, Instituto Nacional de Enfermedades Neoplásicas, and Instituto Nacional de Ciencias Neurológicas. Notably, the Instituto Nacional de Ciencias Neurológicas had half of its articles (50.12%) highly cited (more than 10 citations). Foreign collaboration and publication in Q1 journals were associated with a higher likelihood of being highly cited. Additionally, publishing in English, conducting interventional research, receiving funding, foreign collaboration, and having male authors as last authors increased the likelihood of publication in Q1 journals. Conclusion: The scientific output of specialized institutes in Peru is heterogeneous and growing, although none of the institutes exceeded 100 articles per year. Moreover, collaboration among the institutes is limited. Reference values should be established to enhance the performance of specialized institutes in Peru.

## INTRODUCTION

Health research plays a critical role in identifying, understanding, and proposing solutions to both individual and collective health issues (1). Several institutions contribute to the advancement of health research, among them specialized health institutes, which are essential for a country’s scientific and technological development. These institutes provide vital information for clinical decision-making, guide public health policies, and propose innovative technological solutions (2,3).

In Peru, the organizational structure of the Ministry of Health (MINSA) includes both affiliated public entities focused on research, such as the Instituto Nacional de Salud (INS) and the Instituto Nacional de Enfermedades Neoplásicas (INEN), as well as seven deconcentrated entities, which include specialized institutes like the Instituto Nacional de Ciencias Neurológicas (INCN), Instituto Nacional de Oftalmología (INO), Instituto Nacional de Salud del Niño de San Borja (INSN-SB), Instituto Nacional de Salud del Niño de Breña (INSN-Breña), Instituto Nacional Materno Perinatal (INMP), Instituto Nacional de Salud Mental (INSM), and Instituto Nacional de Rehabilitación (INR) (4–7).

It is important to note that the INS is regarded as a public research institute and the governing body for health research in Peru. Unlike the other institutes, the INS does not provide direct patient care but instead serves as a hub for research activities. The remaining institutes, however, offer highly specialized medical care while engaging in research. Despite these institutions’ mission to foster research, it is concerning that no Peruvian technical regulations set quality standards for research or for evaluating the scientific output of these institutes (8).

Bibliometric studies are valuable tools for assessing scientific performance, as they measure the outcomes of research activity (article production), gauge its dissemination within the academic community through citations (bibliometric impact), and highlight inter-institutional and international collaborations (9). Some bibliometric studies have quantitatively assessed the scientific output of the INS, INSN-Breña, INEN, and others of the institutes mentioned (10–13). However, the bibliometric characteristics of the scientific production across all these institutions, including its temporal evolution, the presence of collaborative networks, and research funding, remain poorly understood.

This study aims to describe the scientific output of specialized health institutes in Peru from 1991 to 2021. Complemented by other management indicators, this information could inform changes in policy and management practices at these institutions. The findings could help decision-makers in the evaluated entities, as well as in similar institutions in middle- and low-income countries, to establish reference baselines for comparing and monitoring scientific productivity.

## METHODS

### Design

A bibliometric study was conducted to analyze the scientific output of specialized health institutes in Peru. The institutes included in the study were: INS, INEN, INCN, INO, INR, INSN-SB, INSN-Breña, INMP, and INSM. The units of analysis were papers published in indexed journals that underwent a peer-review process. Articles included in the study were those written in IMRD format (Introduction, Methods, Results, Discussion), clinical cases, case series, letters to the editor, editorials, special/short communications, narrative reviews (without methodology), symposiums, published between 1991 and 2021, with at least one author affiliated with one of the institutes of interest. The study excluded publications such as ’corrigenda’, ’retracted articles’, preprints, biographies, historical reviews, photographs, conference abstracts, books, and book chapters. Articles that could not be accessed in full were also excluded.

### Databases

The search was carried out in the Web of Science (WoS), Embase, Scopus, and SciELO databases. Advanced search strategies were applied to each database (Embase, WoS, and SciELO), considering the search terms used in Scopus. In the case of SciELO, access was obtained via the WoS search engine using the SciELO Citation Index option. All searches were executed on June 12, 2022. For a detailed description of the search strategies, refer to **Supplementary Material 1**.

### Study selection

Duplicate articles were identified and removed using EndNote, and the accuracy and validity of duplicate results were confirmed using the “duplicates” feature in Microsoft Excel. Some duplicate articles were not detected during the initial filtering process due to variations in language or title formatting. As a result, one author manually identified and removed the remaining duplicates.

### Data extraction

Data extraction was carried out by pairs of researchers, with collaborators trained in research methodology and the critical reading of scientific articles. An induction session was held to explain the methodology and extraction guidelines, using demonstrative examples. Data extraction was performed using a predefined Microsoft Excel® 2021 format. Each article was individually reviewed in its entirety to verify eligibility criteria and extract relevant information on the variables of interest. In case of discrepancies during data extraction, a third author reviewed and resolved the issues.

### Variables

The **general characteristics** evaluated included institutional affiliation, language of publication, medical specialty, funding source, article type, study design, journal name, journal quartile according to Scimago Journal Rank 2022, country of the journal, cross-cutting themes (bioethics, medical education, digital health, participatory medicine), and keywords. Author names and affiliations were manually standardized to account for potential differences in naming conventions. Regarding affiliations, the focus was placed on the main institution, excluding the specific units associated with each institute mentioned. Article type and study design were manually verified for each record by reviewing the full article to classify it as an original publication in IMRD format meeting the relevant criteria. Case reports, letters to the editor, symposiums, special articles, and editorials were grouped under the category of “not applicable.” The medical specialty was assigned based on the article’s title and the specialties of the contributing authors, with a manual standardization allowing for a maximum of three specialties per article.

The **production indicators** included the number of publications per year and across three time periods: 1991-2001, 2002-2011, and 2012-2021, as well as the productivity index by institute and author. The publication year was used as the default year in each database.

The **collaboration indicators** included the co-authorship index, the number of institutional affiliations per article, national and international collaboration rates, and the proportion of authorship leadership affiliated with one of the institutes. The national and international collaboration rate was determined based on the number of authors affiliated with a Peruvian institute, foreign institutions, or both, per article. In cases of multiple affiliations (e.g., two or three types), priority was given to the institute affiliation, followed by Peruvian affiliation, and finally foreign affiliation.

The **bibliometric impact indicators** included the number of citations per year, total citations per institute, citation index per institute, documents ever cited, and highly cited documents (those cited 10 or more times). Citation data were exclusively retrieved from Scopus between February 7th and 9th, 2023. Based on these citations, the CiteScore (total citations divided by the number of citable documents) and the H-index were calculated for each institute globally (1991-2021) and over the past 5 and 10 years.

### Statistical analysis

Categorical data were described using absolute and relative frequencies. For numerical variables, the median and interquartile range were used due to their non-normal distribution, with the mean and standard deviation also provided for comparison. Bibliometric indicators were presented for the entire study period and subperiods (1991-2001, 2002-2011, 2012- 2021), covering production and collaboration, both globally and by institute.

Citation data were extracted from Scopus. Articles from non-indexed journals were labeled as “without citations.” Three scenarios were evaluated: the “Normal case” (excluding “without citations”), and two based on the “best-worst case” model (14). In the best-case scenario (“maximum”), “without citations” were replaced with the highest possible citation count, while in the worst-case (“minimum”), they were replaced with zero.

First authorship was assigned to sole authors or those listed first, while last authorship was assigned to the last author in multi-author articles. A co-occurrence map of institutional collaboration was generated using VOSViewer 4.6.18.

For the thematic analysis, keyword clouds were generated for each institute and globally using WordItOut. A direct color blending method was employed, ranging from green (fewer repetitions) to red (more repetitions), with the color of the words varying (degrading) based on their frequency. Additionally, the size of the words was proportional to their frequency.

The relationship between the variables of interest (highly cited articles and publication in quartile 1) and the associated factors was assessed using the Chi-square test. Adjusted prevalence ratios (PR) with 95% confidence intervals (95% CI) were calculated using Poisson regression with robust variance. The highly cited model was dichotomous (No, Yes) and was associated with the following variables: Language (Spanish, English, Spanish and English, Other), Study type (Basic, Interventional, Not applicable, Observational, Secondary), Journal location (Peru, Latin American, Other), Cross-cutting theme (No, Yes), Funding (No, Yes), Original article (No, Yes), Collaboration (Individual, Peruvian, Foreign), Quartile (No quartile, Q4, Q3, Q2, Q1), First authorship (No, Yes), Gender of first authorship (Female, Male), Last authorship (No, Yes), Gender of last authorship (Female, Male). The publication model in quartile 1 (No, Yes) was associated with the same set of variables, excluding the journal country, as this would be constant since Peruvian and Latin American journals are not present in quartile 1. The analysis was conducted using R software version 4.2.2.

### Ethical aspects

This study followed ethical principles and was approved by the Institutional Ethics Committee of INSN-SB (Code: PI-669, Registration: 029-2022).

## RESULTS

A total of 7,264 articles were identified in the Scopus, WoS, SciELO, and Embase databases. After removing duplicates, 4,413 articles remained for evaluation based on the selection criteria. Ultimately, 3,020 articles were included for formal analysis (see **Figure 1**).

**Figure 1.**
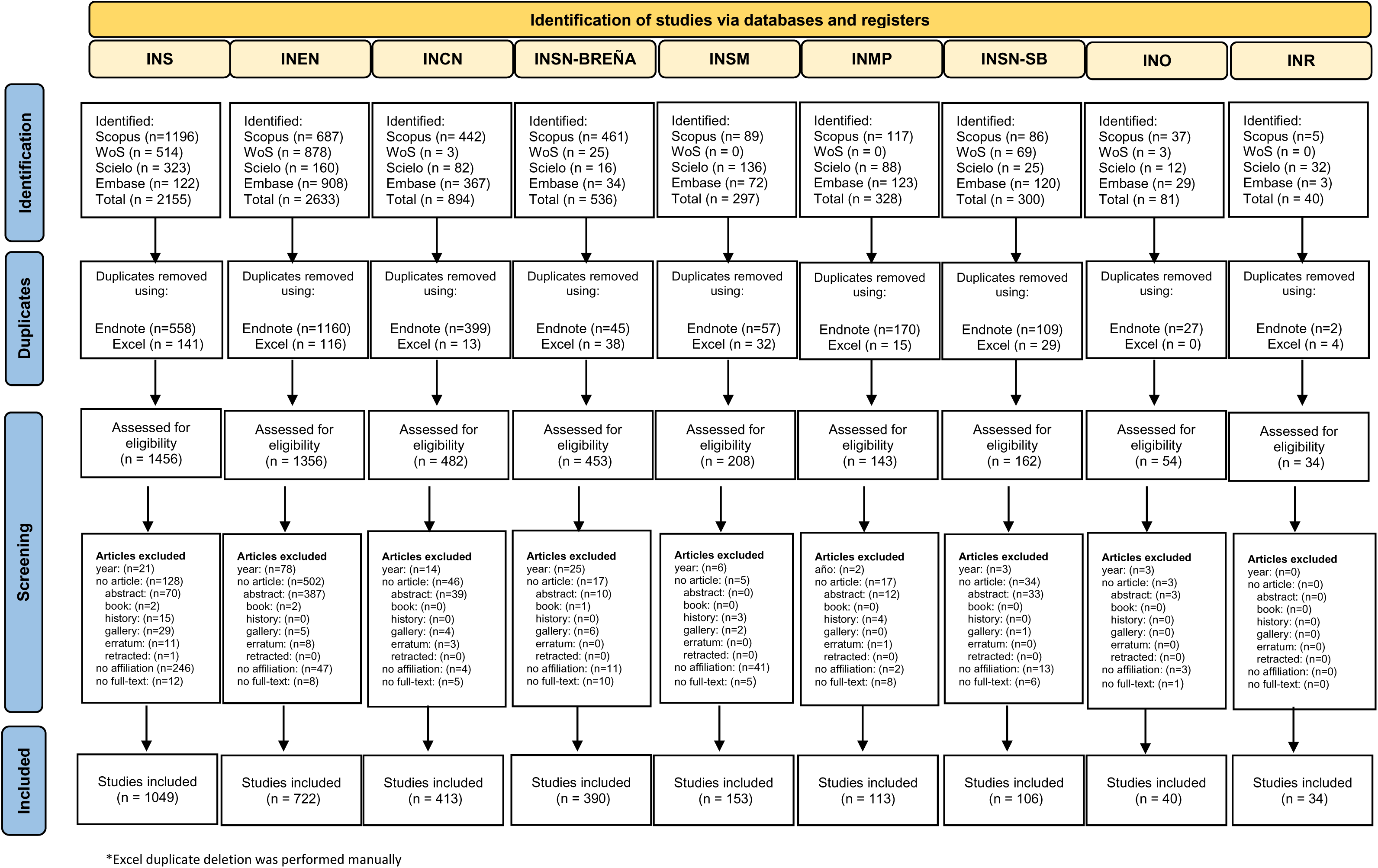
Flowchart summarizing the process of search and study selection of each specialized institute in Peru.

### General characteristics

A total of 3,020 articles from nine institutes were analyzed. The majority of the studies were published in English (61.39%). Among the institutes, INCN had the highest proportion of English publications (85.47%), while INR had the highest proportion of Spanish publications (94.12%). 26.89% of the articles received foreign funding, while 36.92% did not explicitly report receiving any funding. The most frequently published article type across all institutes was original research, and the most common study type was observational. 38.74% of the publications appeared in journals ranked in Q1, with the highest proportion of Q1 publications relative to total output found in INCN (58.11%), INEN (45.71%), INSN-Breña (36.92%), and INSM (18.95%). The second quartile with the highest publication rate was Q3 (27.35%), with INS having the largest proportion of its output in this quartile, while INR had 85.29% of its publications in journals without a quartile ranking. The countries with the highest publication rates in these journals were the USA and European countries, while INSM and INR predominantly published in Peruvian journals. 95.43% of the publications did not address a cross-cutting theme (see **Table 1**).

**Table 1.** General characteristics of scientific output from specialized institutes in Peru.

|  | INS<br>N=1049 | INEN<br>N=722 | INCN<br>N=413 | INSN-Breña<br>N=390 | INSM<br>N=153 | INMP<br>N=113 | INSN-SB<br>N=106 | INO<br>N=40 | INR<br>N=34 | Total<br>N=3020 |
| --- | --- | --- | --- | --- | --- | --- | --- | --- | --- | --- |
| **Language:** |  |  |  |  |  |  |  |  |  |  |
| Spanish | 484 (46.14%) | 194 (26.87%) | 57 (13.80%) | 71 (18.21%) | 93 (60.78%) | 35 (30.97%) | 27 (25.47%) | 16 (40.00%) | 32 (94.12%) | 1009 (33.41%) |
| English | 514 (49.00%) | 518 (71.75%) | 353 (85.47%) | 270 (69.23%) | 50 (32.68%) | 69 (61.06%) | 62 (58.49%) | 17 (42.50%) | 1 (2.94%) | 1854 (61.39%) |
| Spanish and English | 50 (4.77%) | 9 (1.25%) | 3 (0.73%) | 48 (12.31%) | 10 (6.54%) | 8 (7.08%) | 17 (16.04%) | 7 (17.50%) | 1 (2.94%) | 153 (5.07%) |
| Others | 1 (0.10%) | 1 (0.14%) | 0 (0.00%) | 1 (0.26%) | 0 (0.00%) | 1 (0.88%) | 0 (0.00%) | 0 (0.00%) | 0 (0.00%) | 4 (0.13%) |
| **Funding:** |  |  |  |  |  |  |  |  |  |  |
| No mention | 295 (28.12%) | 309 (42.80%) | 125 (30.27%) | 200 (51.28%) | 73 (47.71%) | 39 (34.51%) | 34 (32.08%) | 21 (52.50%) | 19 (55.88%) | 1115 (36.92%) |
| Self-funded | 145 (13.82%) | 154 (21.33%) | 47 (11.38%) | 72 (18.46%) | 49 (32.03%) | 39 (34.51%) | 44 (41.51%) | 9 (22.50%) | 3 (8.82%) | 562 (18.61%) |
| Peruvian and Foreign | 83 (7.91%) | 12 (1.66%) | 35 (8.47%) | 22 (5.64%) | 0 (0.00%) | 0 (0.00%) | 5 (4.72%) | 3 (7.50%) | 0 (0.00%) | 160 (5.30%) |
| Foreign only | 267 (25.45%) | 214 (29.64%) | 195 (47.22%) | 64 (16.41%) | 24 (15.69%) | 27 (23.89%) | 16 (15.09%) | 4 (10.00%) | 1 (2.94%) | 812 (26.89%) |
| Peruvian only | 259 (24.69%) | 33 (4.57%) | 11 (2.66%) | 32 (8.21%) | 7 (4.58%) | 8 (7.08%) | 7 (6.60%) | 3 (7.50%) | 11 (32.35%) | 371 (12.28%) |
| **Article type:** |  |  |  |  |  |  |  |  |  |  |
| Letter to the editor | 162 (15.44%) | 52 (7.20%) | 32 (7.75%) | 27 (6.92%) | 21 (13.73%) | 6 (5.31%) | 13 (12.26%) | 3 (7.50%) | 10 (29.41%) | 326 (10.79%) |
| Case report | 24 (2.29%) | 99 (13.71%) | 47 (11.38%) | 69 (17.69%) | 31 (20.26%) | 15 (13.27%) | 33 (31.13%) | 8 (20.00%) | 8 (23.53%) | 334 (11.06%) |
| Editorial | 45 (4.29%) | 12 (1.66%) | 12 (2.91%) | 8 (2.05%) | 3 (1.96%) | 1 (0.88%) | 1 (0.94%) | 0 (0.00%) | 0 (0.00%) | 82 (2.72%) |
| Original | 714 (68.06%) | 470 (65.10%) | 251 (60.77%) | 258 (66.15%) | 75 (49.02%) | 84 (74.34%) | 51 (48.11%) | 28 (70.00%) | 15 (44.12%) | 1946 (64.44%) |
| Others | 104 (9.91%) | 89 (12.33%) | 71 (17.19%) | 28 (7.18%) | 23 (15.03%) | 7 (6.19%) | 8 (7.55%) | 1 (2.50%) | 1 (2.94%) | 332 (10.99%) |
| **Study type:** |  |  |  |  |  |  |  |  |  |  |
| Basic | 171 (16.30%) | 43 (5.96%) | 85 (20.58%) | 8 (2.05%) | 0 (0.00%) | 1 (0.88%) | 3 (2.83%) | 1 (2.50%) | 0 (0.00%) | 312 (10.33%) |
| Interventional | 27 (2.57%) | 93 (12.88%) | 14 (3.39%) | 26 (6.67%) | 2 (1.31%) | 3 (2.65%) | 5 (4.72%) | 3 (7.50%) | 1 (2.94%) | 174 (5.76%) |
| Not applicable | 314 (29.93%) | 206 (28.53%) | 119 (28.81%) | 126 (32.31%) | 78 (50.98%) | 29 (25.66%) | 53 (50.00%) | 12 (30.00%) | 19 (55.88%) | 956 (31.66%) |
| Observational | 446 (42.52%) | 300 (41.55%) | 126 (30.51%) | 193 (49.49%) | 61 (39.87%) | 73 (64.60%) | 42 (39.62%) | 22 (55.00%) | 14 (41.18%) | 1277 (42.28%) |
| Secondary | 91 (8.67%) | 80 (11.08%) | 69 (16.71%) | 37 (9.49%) | 12 (7.84%) | 7 (6.19%) | 3 (2.83%) | 2 (5.00%) | 0 (0.00%) | 301 (9.97%) |
| **Journal quartile** |  |  |  |  |  |  |  |  |  |  |
| Q1 | 346 (32.98%) | 330 (45.71%) | 240 (58.11%) | 144 (36.92%) | 29 (18.95%) | 42 (37.17%) | 27 (25.47%) | 12 (30.00%) | 0 (0.00%) | 1170 (38.74%) |
| Q2 | 128 (12.20%) | 83 (11.50%) | 66 (15.98%) | 68 (17.44%) | 12 (7.84%) | 14 (12.39%) | 12 (11.32%) | 5 (12.50%) | 1 (2.94%) | 389 (12.88%) |
| Q3 | 438 (41.75%) | 115 (15.93%) | 64 (15.50%) | 116 (29.74%) | 29 (18.95%) | 22 (19.47%) | 34 (32.08%) | 6 (15.00%) | 2 (5.88%) | 826 (27.35%) |
| Q4 | 64 (6.10%) | 116 (16.07%) | 8 (1.94%) | 44 (11.28%) | 10 (6.54%) | 16 (14.16%) | 16 (15.09%) | 12 (30.00%) | 2 (5.88%) | 288 (9.54%) |
| No quartile | 73 (6.96%) | 78 (10.80%) | 35 (8.47%) | 18 (4.62%) | 73 (47.71%) | 19 (16.81%) | 17 (16.04%) | 5 (12.50%) | 29 (85.29%) | 347 (11.49%) |
| **Country of the journal** |  |  |  |  |  |  |  |  |  |  |
| Latin America | 96 (9.15%) | 34 (4.71%) | 10 (2.42%) | 49 (12.56%) | 10 (6.54%) | 13 (11.50%) | 12 (11.32%) | 6 (15.00%) | 1 (2.94%) | 231 (7.65%) |
| USA and Europe | 518 (49.38%) | 515 (71.33%) | 356 (86.20%) | 254 (65.13%) | 56 (36.60%) | 72 (63.72%) | 65 (61.32%) | 24 (60.00%) | 1 (2.94%) | 1861 (61.62%) |
| Peru | 435 (41.47%) | 173 (23.96%) | 47 (11.38%) | 87 (22.31%) | 87 (56.86%) | 28 (24.78%) | 29 (27.36%) | 10 (25.00%) | 32 (94.12%) | 928 (30.73%) |
| **Cross-cutting theme** |  |  |  |  |  |  |  |  |  |  |
| No | 979 (93.33%) | 711 (98.48%) | 400 (96.85%) | 389 (99.74%) | 129 (84.31%) | 109 (96.46%) | 99 (93.40%) | 38 (95.00%) | 28 (82.35%) | 2882 (95.43%) |
| Yes | 70 (6.67%) | 11 (1.52%) | 13 (3.15%) | 1 (0.26%) | 24 (15.69%) | 4 (3.54%) | 7 (6.60%) | 2 (5.00%) | 6 (17.65%) | 138 (4.57%) |

INS: Instituto Nacional de Salud, INEN: Instituto Nacional de Enfermedades Neoplásicas, INCN: Instituto Nacional de Ciencias Neurológicas, INO: Instituto Nacional de Oftalmología, INSN-SB: Instituto Nacional de Salud del Niño de San Borja, INSN-Breña: Instituto Nacional de Salud del Niño de Breña, INMP: Instituto Nacional Materno Perinatal, INSM: Instituto Nacional de Salud Mental, and INR: Instituto Nacional de Rehabilitación.

Regarding the funding of original articles, INCN had the highest percentage of foreign funding (62.55%). Despite its lower total output, INR had a significant proportion of its original articles funded by Peruvian sources (see **Supplementary Material 2**). The journal with the highest number of publications from specialized institutes in Peru was *Revista Peruana de Medicina Experimental y Salud Pública* (Q3), which accounted for 510 articles, or 16.89% of the institutes’ total production. INS published 33.74% of its articles in this journal (see **Supplementary Material 3**). The most researched specialty was “Medicine of Infectious and Tropical Diseases,” which accounted for one-third (31.03%) of the institutes’ total production (see **Supplementary Material 4**). In terms of keyword analysis, the most frequent terms were “Peru” and “health,” followed by “cancer” and “tuberculosis” (see **Supplementary Material 5**).

### Productivity indicators

The institute with the highest scientific production was INS, with 1,049 publications, followed by INEN with 722. However, in the last year (2021), INEN published more articles than INS (96 vs. 81) (see **Figure 2**). In the analysis of subperiods of 10 years, INSN-SB showed increased productivity in the last decade, which corresponds to its recent creation in 2013. In contrast, INR (1962) and INO (1987), which are older, have lower total scientific production (see **Supplementary Material 6**). It is noteworthy that INCN, INSN-SB, INSN- Breña, and INS were the only institutes showing a positive productivity growth rate between 2020 and 2021 (74.19%, 56%, 27.45%, and 3.85%, respectively). INEN was the only institute with no change in production during this period. Interestingly, the three most productive authors of INCN contributed to more than three-quarters of its total output (77.24%). In terms of gender-based productivity indices, INR was the only institute with a higher prevalence of first and last female authorship (1.31 and 1.57, respectively) (see **Supplementary Material 7**).

**Figure 2.**
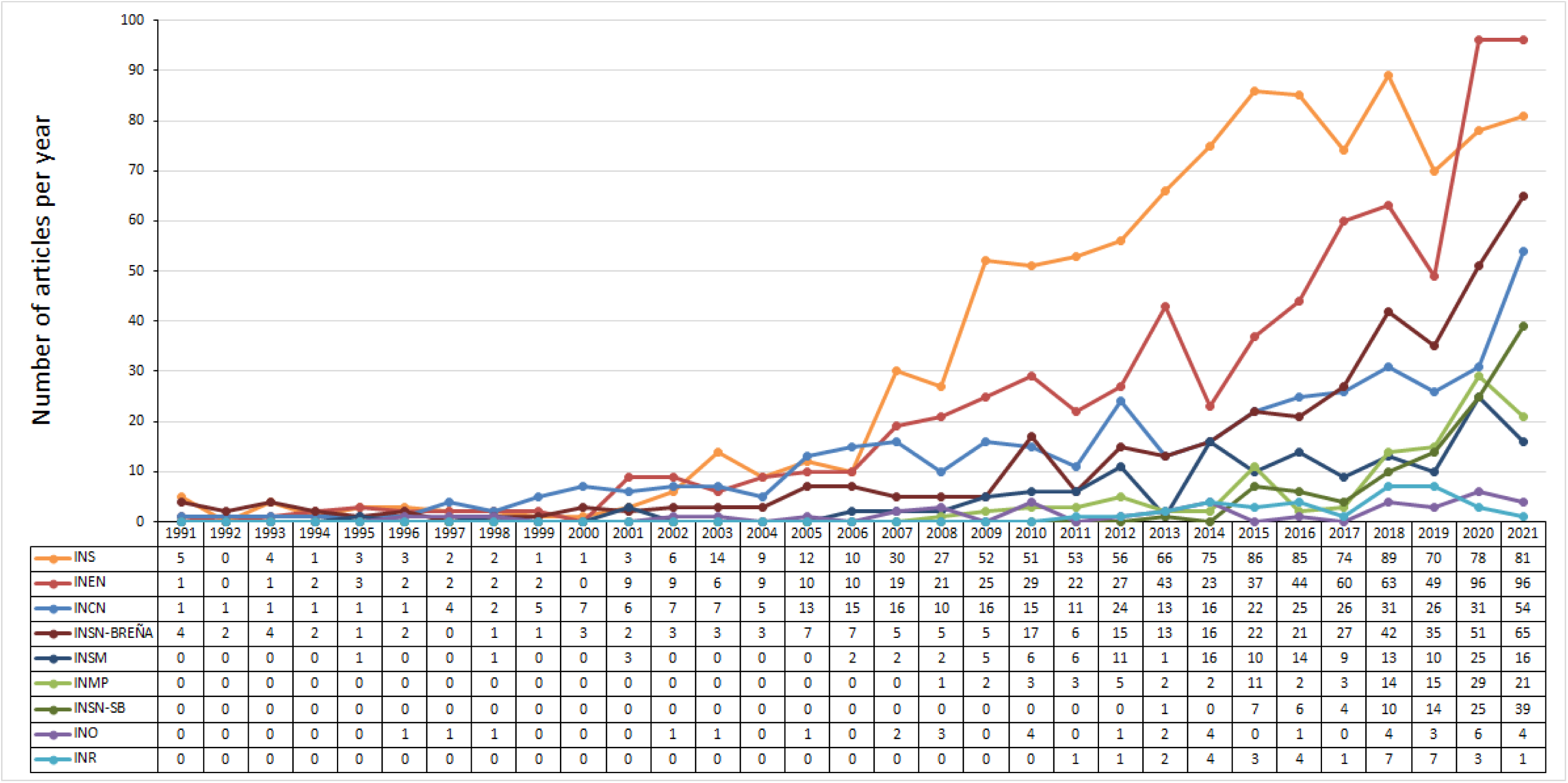
The scientific output of specialized institutes in Peru between 1991 - 2021.

### Collaboration indicators

INS was the institute with the most inter-institute collaboration. On the other hand, INO had the least collaboration with other institutes (see **Supplementary Material 8**). The institute with the highest median number of authors per document was INEN (8; RIC = 5 - 14), followed by INCN and INSN-Breña (both with 7; RIC = 4 - 11). INEN and INCN had a median of two authors per paper from their institute (2; RIC = 1 - 4 and 2; RIC = 1 - 3, respectively), similar to INMP, INO, and INR (2; RIC = 1 - 3, 2; RIC = 1 - 3.25, and 1.5; RIC = 1 - 3, respectively), despite having fewer articles. Notably, INEN, INSM, and INR had medians of zero Peruvian authors per document (RIC = 0 - 1; RIC = 0 - 1; RIC = 0 - 0, respectively), while INEN had a median of 1 (RIC = 0 - 11) for the number of foreign authors. This suggests that for INEN, 50% of the articles had between 0 and 1 Peruvian authors, and 50% had between 0 and 11 foreign authors per document (see **Table 2**). The institute with the highest foreign collaboration was INEN (41.97%), while INCN had the highest percentage of both foreign and Peruvian collaboration (40%). INR was the institute with the highest percentage of solo institutional production (74%) (see **Figure 3**).

**Figure 3.**
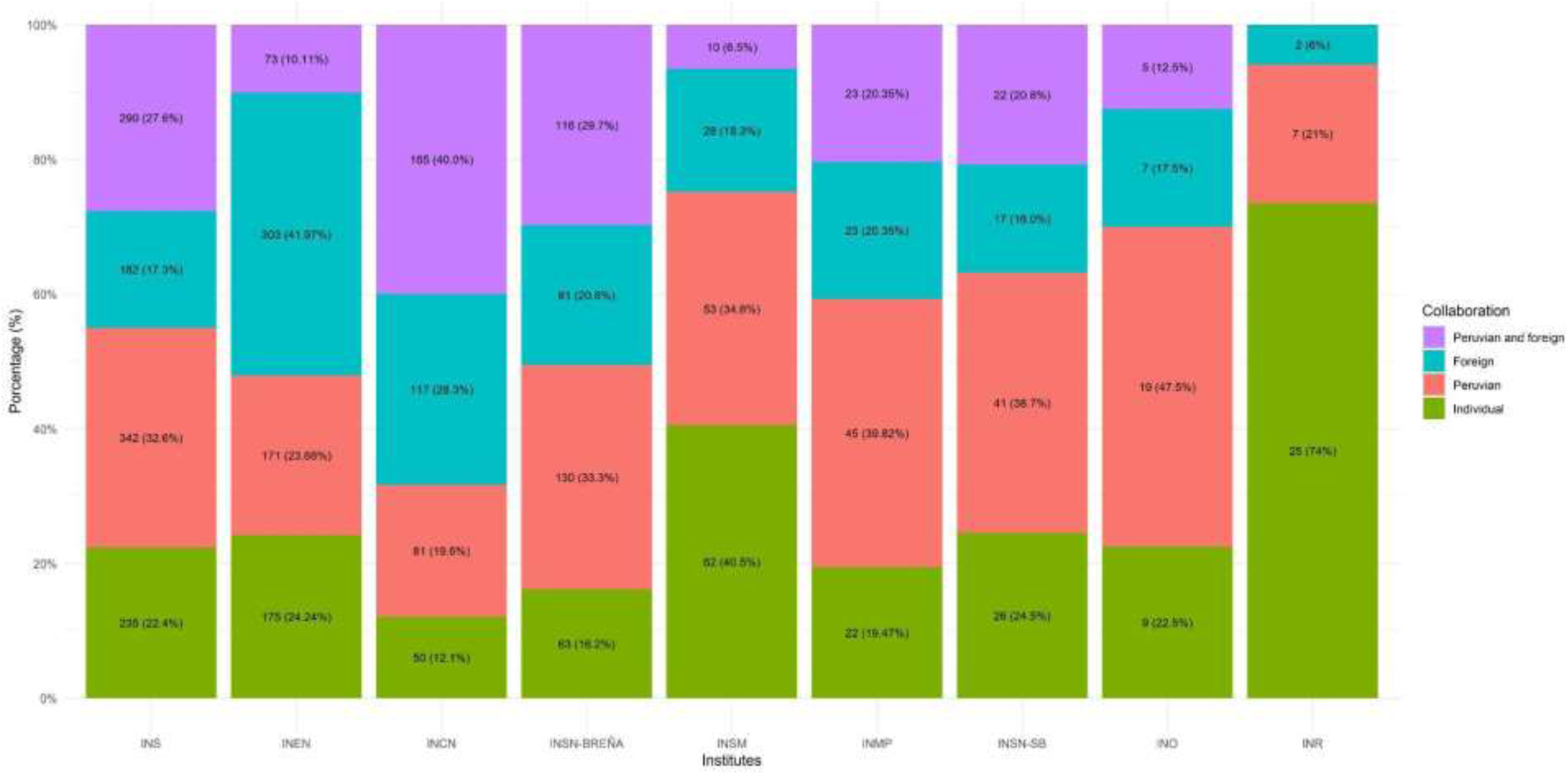
National and international collaboration by specialized institutes of health in Peru.

**Table 2.** Means and medians of authors, both national and international, by institute.

|  | **INS**<br>**(N=1049)** | **INEN**<br>**(N=722)** | **INCN**<br>**(N=413)** | **INSN-Breña**<br>**(N=390)** | **INSM**<br>**(N=153)** | **INMP**<br>**(N=113)** | **INSN-SB**<br>**(N=106)** | **INO**<br>**(N=40)** | **INR**<br>**(N=34)** | **Total**<br>**(N=3020)** |
| --- | --- | --- | --- | --- | --- | --- | --- | --- | --- | --- |
| **Number of authors per document** |  |  |  |  |  |  |  |  |  |  |
| Mean (SD) | 12.5 (33.2) | 12.3 (18.4) | 9.13 (10.2) | 9.22 (9.32) | 5.84 (7.66) | 12.1 (29.2) | 12.0 (23.2) | 5.78 (3.33) | 2.68 (1.79) | 11.0 (23.4) |
| Median [IQR] | 6 [3;10] | 8 [5;14] | 7 [4;11] | 7 [4;11] | 3 [2;5] | 6 [4;9] | 6 [4;11] | 5 [4;7] | 2 [1;4] | 6 [4;11] |
| **Number of authors per document from Institutes** |  |  |  |  |  |  |  |  |  |  |
| Mean (SD) | 2.21 (1.96) | 2.93 (3.01) | 2.43 (2.08) | 2.12 (2.01) | 1.69 (1.11) | 2.66 (2.69) | 2.65 (2.46) | 2.45 (1.68) | 2.12 (1.43) | 2.41 (2.31) |
| Median [IQR] | 1 [1;3] | 2 [1;4] | 2 [1;3] | 1 [1;2] | 1 [1;2] | 2 [1;3] | 1 [1;4] | 2 [1;3.25] | 1.5 [1;3] | 1 [1;3] |
| **Number of authors per document from Peru** |  |  |  |  |  |  |  |  |  |  |
| Mean (SD) | 1.79 (2.39) | 1.13 (2.27) | 2.59 (3.53) | 2.52 (3.37) | 0.96 (1.95) | 3.70 (19.0) | 2.58 (3.41) | 2.20 (3.12) | 0.38 (1.02) | 1.88 (4.58) |
| Median [IQR] | 1 [0;3] | 0 [0;1] | 1 [0;4] | 2 [0;4] | 0 [0;1] | 1 [0;3] | 1 [0;4] | 1 [0;3.25] | 0 [0;0] | 1 [0;3] |
| **Number of authors per document Foreign** |  |  |  |  |  |  |  |  |  |  |
| Mean (SD) | 8.56 (33.7) | 8.22 (18.1) | 4.11 (9.76) | 4.56 (9.63) | 3.18 (7.99) | 5.72 (22.9) | 6.79 (23.5) | 1.12 (2.13) | 0.18 (0.87) | 6.72 (23.3) |
| Median [IQR] | 0 [0;5] | 1 [0;11] | 2 [0;5] | 1 [0;5] | 0 [0;0] | 0 [0;5] | 0 [0;2] | 0 [0;2] | 0 [0;0] | 0 [0;5.75] |

SD: Standard deviation, IQR: Interquartile range

Regarding leadership, of the 3,020 articles, 1,712 (56.69%) had first authorship not affiliated with specialized institutes. Despite its lower production, INR had the highest percentage of first authorship leadership (88.2%), followed by INSM and INSN-SB (64.1% and 51.9%, respectively). INR was the only institute with a higher percentage of female first authorship (56.7%) (see **Supplementary Material 9**).

### Bibliometric impact indicators

The institute with the highest median citations per document was INCN (12; IQR = 4 - 34), followed by INS, INEN, and INO (all with a median of 7; RIC = 2 - 22, RIC = 2 - 28.5, and RIC = 0 - 12.5, respectively). This indicates that 50% of INCN articles received between 4 and 34 citations. INCN demonstrated significant impact, as 50.12% of its articles were highly cited, and 353 (85.47%) had at least one citation. Despite a higher total publication count, only 37.85% (397) of INS articles were highly cited, though 812 (77.41%) were ever cited. INO, despite its smaller output, had one-third of its articles highly cited (32.5%), compared to larger institutes like INMP, INSM, and INSN-SB (25.66%, 22.22%, and 16.04%, respectively). INS had the highest CiteScore (64.96) and H-index (76). Notably, INO had a higher CiteScore but a lower H-index than INSN-SB (11.94 vs. 5.59, and 11 vs. 12, respectively) (see **Table 3**).

**Table 3.** Impact indicators for each specialized institute in Peru.

|  | INS | INEN | INCN | INSN-Breña | INSM | INMP | INSN-SB | INO | INR |
| --- | --- | --- | --- | --- | --- | --- | --- | --- | --- |
| **Total number of articles (T)** | 1049 | 722 | 413 | 390 | 153 | 113 | 106 | 40 | 34 |
| **Total number of citations per institute** | 59890 | 28038 | 11872 | 7649 | 2061 | 1118 | 503 | 370 | 23 |
| **Number of citations by document per institute** |  |  |  |  |  |  |  |  |  |
| Minimum |  |  |  |  |  |  |  |  |  |
| Mean (SD) | 57.1 (345) | 38.8 (125) | 28.7 (58.9) | 19.6 (62.2) | 13.5 (42.2) | 9.89 (18.8) | 4.75 (8.64) | 9.25 (17.7) | 0.68 (2.45) |
| Median [IQR] | 5 [1;17] | 5 [0;23] | 10 [3;32] | 4 [0;15.8] | 0 [0;5] | 3 [0;12] | 1 [0;5] | 2 [0;12] | 0 [0; 0] |
| Normal |  |  |  |  |  |  |  |  |  |
| Mean (SD) | 65.0 (367) | 45.9 (134) | 31.3 (60.9) | 20.2 (63.1) | 26.1 (56.0) | 12.2 (20.2) | 5.59 (9.13) | 11.9 (19.3) | 4.60 (5.18) |
| Median [IQR] | 7 [2;22] | 7 [2;28.5] | 12 [4;34] | 4 [1;16] | 5 [1;17] | 4.5 [1;14] | 1.5 [0;6] | 7 [0;12.5] | 2 [2;6] |
| Maximum |  |  |  |  |  |  |  |  |  |
| Mean (SD) | 772 (1938) | 319 (653) | 81.9 (179) | 45.4 (154) | 161 (145) | 28.7 (39.2) | 11.5 (16.5) | 30.9 (39.4) | 11.8 (3.52) |
| Median [IQR] | 9 [2;42] | 12 [3;86.8] | 15 [4;45] | 5 [1;20.8] | 197 [4;304] | 9 [2;36] | 2.5 [0.25;14.5] | 11.5 [1.75;57] | 13 [13;13] |
| **Number of citations of the most cited article** | 5908 | 1823 | 646 | 837 | 304 | 101 | 45 | 96 | 13 |
| **Number of highly cited publications (X)** | 397 | 281 | 207 | 138 | 34 | 29 | 17 | 13 | 1 |
| **Percentage of highly cited publications (highly cited) (X/T)** | 37,85 | 38,97 | 50,12 | 35,38 | 22,22 | 25,66 | 16,04 | 32,50 | 2,94 |
| **Number of publications ever cited (Y)** | 812 | 521 | 353 | 291 | 73 | 63 | 62 | 22 | 4 |
| **Percentage of documents ever cited (Y/T)** | 77,41 | 72,26 | 85,47 | 74,62 | 47,71 | 55,75 | 58,49 | 55,00 | 11,76 |
| **Total citations in Scopus\* (a)** | 59890 | 28038 | 11872 | 7649 | 2061 | 1118 | 503 | 370 | 23 |
| **Scopus citable papers\* (b)** | 922 | 611 | 379 | 378 | 79 | 92 | 90 | 31 | 5 |
| **Cite score\* (a/b)** | 64.96 | 45.89 | 31.32 | 20.24 | 26.09 | 12.15 | 5.59 | 11.94 | 4.6 |
| **H-index (1991-2021)\*** | 76 | 74 | 56 | 43 | 18 | 18 | 12 | 11 | 2 |
| **H5-index (2017 - 2021)\*** | 42 | 37 | 18 | 19 | 7 | 12 | 10 | 3 | 1 |
| **H10-index (2012 - 2021)\*** | 69 | 53 | 33 | 29 | 11 | 16 | 12 | 6 | 2 |

SD: Standard deviation, IQR: Interquartile range, \*: Calculation made with data extracted from bibliometric citation data.
Note: Only papers that had a Scopus citation were considered.

The “highly cited” variable showed positive associations with publishing in quartile 1 (PRa = 2.06; 95% CI = 1.15 - 3.66), foreign collaboration (PRa = 1.42; 95% CI = 1.1 - 1.84), and male first authorship (PRa = 1.11; 95% CI = 1.02 - 1.2). Additionally, “publication in quartile 1” was positively associated with publishing in English (PRa = 71.92; 95% CI = 29.6 - 174.74), foreign collaboration (PRa = 1.27; 95% CI = 1.01 - 1.6), funding (PRa = 1.23; 95% CI = 1.13 - 1.34), interventional study type (PRa = 1.17; 95% CI = 1.05 - 1.3), and male last authorship (PRa = 1.16; 95% CI = 1.06 - 1.28) (see **Supplementary Material 10**).

## DISCUSSION

### Production indicators

The analysis revealed that the institutes published more articles in the most recent period (2012–2021), confirming the sustained growth of Peruvian scientific production in recent years (15). However, it is important to note that these institutes have not yet reached the minimum of 100 articles required to be considered in the SCImago Ranking of Institutions (16). Variations in production could be attributed to factors such as the institutes’ age (older institutes, like INS and INEN, publish more) (17,18) and the priority given to research by leadership. While most institutes mention research in their missions, only INS and INEN include it in their strategic plans (19–25). Other factors include research management, R&D funding (26), infrastructure, human capital, and collaborative networks. The decline or stagnation in the number of publications in 2021, observed in half of the institutes, can likely be attributed to the COVID-19 pandemic. Many institutions had to shift their focus to COVID-19-related research and reorganize staff and resources to address the health crisis 27). Recently, the Peruvian government established performance indicators and improvement commitments for services to be met in 2023, requiring specialized institutes to publish more than five original articles per year to achieve the maximum score (28,29). While this goal is achievable, it may not significantly boost research activity, indicating a lack of emphasis on research promotion by the government.

### Collaboration indicators

It was found that INR, INSM, and INSN-SB had the highest percentages of leadership, but at the same time, they also had the highest percentages of publications without collaboration (74%, 40.5%, and 24.5%, respectively). These findings, regarding the association between high leadership and low levels of collaboration, align with a previous study of health institutes in Cuba (30). It is well-established that high leadership is a merit when associated with a high rate of collaboration (especially international collaboration), as it generally leads to greater visibility and citation (30,31). This aligns with our findings, as INR shows the highest leadership but a very low percentage of international collaboration (5.88%) and highly cited publications (2.94%). Similarly, INCN does not have high leadership (first authorship: 38.5%, last authorship: 37%), but it has the second-highest percentage of foreign collaboration (28.3%) and the highest percentage of highly cited publications (50.12%). These results are significant as they highlight the importance of Peruvian research collaboration with foreign countries to enhance visibility and citation, in addition to providing valuable learning opportunities and improving the quality of research. However, it should be noted that in collaborations between low- and middle-income countries and high-income countries, there may be various gaps, such as those related to language, power, and prestige. These gaps could lead to inequities in the distribution of responsibilities and rewards (e.g., authorship order) (32,33). This underscores the importance of promoting both leadership and collaboration.

### Bibliometric impact indicators

The three institutes with the highest bibliometric impact were INS, INEN, and INCN. INCN stands out for having half of its articles highly cited, indicating strong global visibility. The likelihood of an article being highly cited is associated with factors such as publishing in quartile 1 journals and foreign collaboration, as demonstrated in this study and a previous report by Yaminfirooz et al. (34), which also linked citation rates to the H-index and the number of papers by the first author.

A total of 61.39% of the studies were published in English, which could be attributed to the authors’ interest in disseminating their results internationally, with the goal of reaching a broader global audience and increasing their likelihood of being cited (35,36). Although this study did not find evidence that publishing in English directly increases the likelihood of an article being highly cited, it did find that it increases the probability of publication in a journal positioned in quartile 1.

Beyond the intention of achieving a higher number of citations through language choice, it is important to consider that the research findings may not be accessible to the target population (Peruvians) due to the language barrier, as English is not their native language (37). One possible solution to this issue is selecting journals that publish articles in both English and Spanish. Alternatively, researchers could share their articles by providing translations as supplementary material in the chosen journal or through web platforms such as Zenodo, ResearchGate, or Academia.edu.

### Strengths

This study adopted a broad frame of reference, both in terms of databases and time, allowing for a comprehensive visualization of the evolution of the institutes over the years. Additionally, key variables related to general characteristics, production, collaboration, and impact were considered to provide a thorough characterization of the scientific output of specialized institutes in Peru. Moreover, an association analysis was conducted—an approach rarely employed in bibliometric studies—to explore the relationship between highly cited publications and publication in quartile 1 journals, which will help direct future efforts toward these variables. Notably, this is the first study to assess the scientific productivity of specialized institutions across all of Peru, a middle-income country, in an area that has been largely unexplored and lacks evidence. It is also important to highlight that this study includes a historical comparative analysis (1991–2021) of nine specialized institutions nationwide. This research lays the foundation for the evaluation and performance of national public institutions in the health sector, offering a clear overview of expectations for health institutions regarding research and setting objectives, strategies, and realistic indicators for the years to come.

### Limitations

Bibliometric studies focus on the quantity of published research and its dissemination through citations, without assessing research quality or its broader impact. As a result, the conclusions of this study are limited in scope but provide a useful evaluation of the scientific performance of specialized health institutes in Peru using internationally recognized indicators.

## Conclusion

The scientific output of specialized institutes in Peru is growing but remains heterogeneous, with none exceeding 100 articles per year. While there is significant international collaboration, inter-institutional collaboration is limited. INCN, INEN, and INS stand out for having the highest number of highly cited publications, indicating greater global visibility. However, the lack of appropriate indicators hinders the development of effective strategies to improve research in the country. It is crucial to develop indicators that assess not only the quantity but also the quality of research, in line with principles outlined in the San Francisco Declaration on Research Assessment (DORA) (38). Additionally, measuring impact beyond citations, considering socioeconomic changes, and encouraging interdisciplinary collaboration are essential (39, 40). Evaluating the performance of other healthcare institutions, both public and private, is also necessary. These efforts are key to positioning research as the foundation for technological development and innovation, particularly in developing countries like Peru, where science can drive socioeconomic progress.

## Authors’ contributions

FBC, FZC, and CQJ conceived and designed the research; FBC, FZC, JMV, ESO, AMA, BLM, EV, and CQV collected the data; FBC and FZC were responsible for data curation; FZC analyzed the data; FBC, FZC, and CQJ supervised the research; all authors contributed to and approved the final version of the article. All authors take responsibility for the content of the article and are committed to adequately addressing any questions that may arise to ensure the accuracy of the data and the integrity of any part of their research.

## AI and AI-Assisted Technologies Statement

ChatGPT 4o mini was used exclusively to refine the English writing style.

## Acknowledgments

Thanks to Shalom Aperrigue, Bruno Olivera, Luis Cutipa Condori, Salvador Tejada, Yanisa Zela-Coila, and Ninoska Cuentas for their contribution as collaborators in the data extraction process

## Data availability statement

All data produced in the present study are available upon reasonable request to the authors

