## Supplementary1-10 for "Uncovering research patterns of a middle-income country: A Comprehensive Study of Peru’s Specialized Health Institutes’ Scientific Output (1991-2021)"

**Supplementary Material 1.** Search strategies in Scopus, WoS, SciELO, and Embase.

| # | Institute | Search strategy |
|---|---|---|
| SCOPUS SEARCH | | |
| #1 | Instituto Nacional de Salud | AF-ID("Instituto Nacional de Salud, Lima" 60071247) OR ( AFFIL ( "Instituto Nacional de Salud, Lima" ) OR AFFIL ( "Instituto Nacional De Salud" W/5 ( lima OR peru ) ) OR AFFIL ( "Instituto Nacional De Salud.Lima" ) OR AFFIL ( "Instituto Nacional De Salud (ins)" W/5 ( lima OR peru ) ) OR AFFIL ( "Instituto National De Salud" W/5 ( lima OR peru ) ) OR AFFIL ( "National Institute Of Health" W/5 ( lima OR peru ) ) OR AFFIL ( "Instituto Nacional De Salud Del Peru" ) OR AFFIL ( "INS" W/5 ( lima OR peru ) ) ) |
| #2 | Instituto Nacional de Enfermedades Neoplásicas | AF-ID("Instituto Nacional de Enfermedades Neoplásicas" 60071241) OR ( AFFIL ( "Instituto Nacional de Enfermedades Neoplásicas" W/5 ( lima OR peru ) ) OR AFFIL ( "Instituto Nacional De Enfermedades Neoplasicas" W/5 ( lima OR peru ) ) OR AFFIL ( "Instituto Nacional De Enfermedades Neoplásicas (inen)" W/5 ( lima OR peru ) ) OR AFFIL ( "National Institute Of Neoplastic Diseases" W/5 ( lima OR peru ) ) OR AFFIL ( "Instituto Nacional De Enfermedades Neoplásicas. Lima" ) OR AFFIL ( "Instituto De Enfermedades Neopl( AFFIL ( "Instituto Nacional de Ciencias Neurológicas" W/5 ( lima OR peru ) ) OR AFFIL ( "Instituto Nacional De Ciencias Neurológicas" W/5 ( lima OR peru ) ) OR AFFIL ( "Instituto Nacional De Ciencias Neurologicas" W/5 ( lima OR peru ) ) OR AFFIL ( "Instituto De Ciencias Neurologicas" W/5 ( lima OR peru ) ) OR AFFIL ( "Instituto De Ciencias Neurológicas" W/5 ( lima OR peru ) ) OR AFFIL ( "National Institute Of Neurological Sciences" W/5 ( lima OR peru ) ) OR AFFIL ( "Inst. Nac. De Ciencias Neurologicas" W/5 ( lima OR peru ) ) OR AFFIL ( "Inst Especializado En Cie. Neurol." W/5 ( lima OR peru ) ) OR AFFIL ( "Instituto Especializado En Ciencias Neurológicas" W/5 ( lima OR peru ) ) OR AFFIL ( "Inst. Nac. De Cie. Neurologicas" W/5 ( lima OR peru ) ) OR AFFIL ( "Inst. De Cie. Neurológicas" W/5 ( lima OR peru ) ) )ásicas" W/5 ( lima OR peru ) ) OR AFFIL ( "Instituto De Enfermedades Neoplasicas" W/5 ( lima OR peru ) ) OR AFFIL ( "Instituto Nacional De Enfermedades Neoplasicas (inen)" ) OR AFFIL ( "INEN" W/5 ( lima OR peru ) ) ) |
| #3 | Instituto Nacional de Ciencias Neurológicas | AF-ID ( "Instituto Nacional de Ciencias Neurologicas" 60071243 ) OR ( AFFIL ( "Instituto Nacional de Ciencias Neurológicas" W/6 ( lima OR peru ) ) OR AFFIL ( "Instituto Nacional De Ciencias Neurológicas" W/6 ( lima OR peru ) ) OR AFFIL ( "Instituto Nacional De Ciencias Neurologicas" W/6 ( lima OR peru ) ) OR AFFIL ( "Instituto De Ciencias Neurologicas" W/6 ( lima OR peru ) ) OR AFFIL ( "Instituto De Ciencias Neurológicas" W/6 ( lima OR peru ) ) OR AFFIL ( "National Institute Of Neurological Sciences" W/6 ( lima OR peru ) ) OR AFFIL ( "Inst. Nac. De Ciencias Neurologicas" W/6 ( lima OR peru ) ) OR AFFIL ( "Inst Especializado En Cie. Neurol." W/6 ( lima OR peru ) ) OR AFFIL ( "Instituto Especializado En Ciencias Neurológicas" W/6 ( lima OR peru ) ) OR AFFIL ( "Inst. Nac. De Cie. Neurologicas" W/6 ( lima OR peru ) ) OR AFFIL ( "Inst. De Cie. Neurológicas" W/6 ( lima OR peru ) ) OR AFFIL ( "INCN" W/6 ( lima OR peru ) ) ) |
| #4 | Instituto Nacional de | AF-ID ( "Instituto Nacional de Oftalmología "Dr. Francisco Contreras Campos"" 60117256 ) OR ( AFFIL ( instituto AND |

| | | |
|---|---|---|
| | Oftalmología | nacional AND de AND oftalmología AND "Dr. Francisco Contreras Campos" ) OR AFFIL ( "Instituto Nacional De Oftalmología" W/5 ( lima OR peru ) ) OR AFFIL ( "Instituto Nacional De Oftalmología Dr. Francisco Contreras Campos" ) OR AFFIL ( "Instituto Nacional De Oftalmologia" W/5 ( lima OR peru ) ) OR AFFIL ( "National Institute Of Ophthalmology" W/5 ( lima OR peru ) ) OR AFFIL ( "Instituto De Oftalmología" W/5 ( lima OR peru ) ) OR AFFIL ( "Instituto De Oftalmologia" W/5 ( lima OR peru ) ) OR AFFIL ( instituto AND nacional AND de AND oftalmología AND "dr. Francisco Contreras Campos" ) OR AFFIL ( "National Institute Of Ophthalmology Of Peru" ) OR AFFIL ( instituto AND nacional AND de AND oftalmología AND "francisco Contreras Campos" ) OR AFFIL ( instituto AND nacional AND de AND oftalmología AND "francisco Contreras Campos," ) OR AFFIL ( "INO" W/5 ( lima OR peru ) ) ) |
| #5 | Instituto Nacional de Rehabilitación | ( AFFIL ( "Instituto Nacional de Rehabilitación" W/5 ( lima OR peru ) ) OR AFFIL ( "Instituto Nacional de Rehabilitacion dra. Adriana Rebaza Flores Amistad Peru Japon" W/5 ( lima OR peru ) ) OR AFFIL ( "Instituto de Rehabilitacion" W/5 ( lima OR peru ) ) OR AFFIL ( "INR" W/5 ( lima OR peru ) ) ) |
| #6 | Instituto Nacional de Salud del Niño de San Borja | AF-ID(“Instituto Nacional de Salud del Niño de San Borja” 60112690) OR ( AFFIL ( "Instituto Nacional de Salud del Niño de San Borja" ) OR AFFIL ( "Instituto Nacional De Salud Del Niño San Borja" ) OR AFFIL ( "Instituto Nacional De Salud Del Niño-san Borja" ) OR AFFIL ( "Instituto Nacional De Salud Del Niño De San Borja" ) OR AFFIL ( "Instituto Nacional De Salud Del Niño San Borja (insn-sb)" ) OR AFFIL ( "Instituto Nacional De Salud Del Niño -- San Borja" ) OR AFFIL ( "Instituto Nacional De Salud Del Niño - San Borja" ) OR AFFIL ( "Instituto Nacional De Salud Del Nino San Borja" ) OR AFFIL ( "Cuidados Paliativos Del Instituto Nacional De Salud Del Niño De San Borja" ) OR AFFIL ( "Instituto Nacional Del Niño San Borja" ) OR AFFIL ( "Instituto Nacional Del Niño" W/5 ( sb OR "San Borja" OR lima OR peru ) ) OR AFFIL ( "insn-sb" ) ). |
| #7 | Instituto Nacional de Salud del Niño de Breña | AF-ID (“Instituto Nacional de Salud del Niño de Breña” 60071245) OR ( AFFIL ( "Instituto Nacional de Salud del Niño de Breña" ) OR AFFIL ( "Instituto Nacional De Salud Del Niño" ) OR AFFIL ( "National Institute Of Child Health" W/6 ( lima OR peru OR breña ) ) OR AFFIL ( "Instituto De Salud Del Niño" ) OR AFFIL ( "Instituto Nacional De Salud Del Niño breña" ) OR AFFIL ( "Instituto Nacional De Salud Del Nino, Lima" ) OR AFFIL ( "Instituto Nacional De Salud Del Nino" ) OR AFFIL ( "Instituto Nacional De Salud Del Niño Lima" ) OR AFFIL ( "Instituto Nacional De Salud Del Niño Breña" ) OR AFFIL ( "Instituto De Salud Del Nino Hosp Nino" ) ) |
| #8 | Instituto Nacional Materno Perinatal | AF-ID ( "Instituto Nacional Materno Perinatal Lima" 60105299 ) OR ( AFFIL ( "Instituto Nacional Materno Perinatal" W/5 ( lima OR peru ) ) OR AFFIL ( "Instituto Nacional Materno Perinatal" W/5 ( lima OR peru ) ) OR AFFIL ( "Instituto Nacional Materno Perinatal De Lima" ) OR AFFIL ( "Instituto Nacional Materno-perinatal" ) OR AFFIL ( "Instituto Nacional Materno Perinatal Inmp" ) OR AFFIL ( "Universidad Peruana Cayetano Heredia And Instituto Nacional Materno Perinatal" ) OR AFFIL ( "Servicio De Medicina Fetal Del Instituto Nacional Materno Perinatal" W/5 ( lima OR peru ) ) OR AFFIL ( "Servicio De Cuidados Intensivos Del Instituto Nacional Materno Perinatal" W/5 ( lima OR peru ) ) OR AFFIL ( "National Maternal Perinatal Institute" W/5 ( lima OR peru ) ) OR AFFIL ( "National Maternal Perinatal Institute" W/5 ( lima OR peru ) ) OR AFFIL ( "National Institute For Maternal And Perinatal Health (inmp)" W/5 ( lima OR peru ) ) OR AFFIL ( "Instituto Nacional Materno Perinatal Universidad Privada Del Norte Lima" ) OR AFFIL ( "INMP" W/5 ( lima OR peru ) ) ) |

| | | |
|---|---|---|
| #9 | Instituto Nacional de Salud Mental | AF-ID(“Instituto Nacional de Salud Mental Honorio Delgado-Hideyo Noguchi” 60103701) OR ( AFFIL ( "Instituto Nacional de Salud Mental Honorio Delgado-Hideyo Noguchi" ) OR AFFIL ( "National Institute Of Mental Health Honorio Delgado Hideyo Noguchi" ) OR AFFIL ( "National Institute Of Mental Health honorio Delgado Hideyo Noguchi" ) OR AFFIL ( "Peruvian National Institute Of Mental Health" W/5 ( lima OR peru ) ) OR AFFIL ( "National Institute Of Mental Health" W/5 ( lima OR peru ) ) OR AFFIL ( "Instituto Nacional De Salud Mental honorio Delgado - Hideyo Noguchi" ) OR AFFIL ( "Instituto Nacional De Salud Mental honorio Delgado-hideyo Noguchi" ) OR AFFIL ( "Instituto Nacional De Salud Mental honorio Delgado-hideyo Noguchi" ) OR AFFIL ( "Instituto Nacional De Salud Mental honorio Delgado -- Hideyo Noguchi" ) OR AFFIL ( "Instituto Nacional De Salud Mental Honorio Delgado Hideyo Noguchi" ) OR AFFIL ( "Instituto Nacional de Salud Mental" W/5 ( lima OR peru ) ) OR AFFIL ( "INSM" W/5 ( lima OR peru ) ) ) |
| WOS SEARCH | | |
| #1 | Instituto Nacional de Salud | (OG=(INS Peru)) OR OG=(Instituto Nacional de Salud - Peru) **OR** (OO=(INS Peru)) OR OO=(Instituto Nacional de Salud - Peru) **OR** (AD=(INS Peru)) OR AD=(Instituto Nacional de Salud - Peru) |
| #2 | Instituto Nacional de Enfermedades Neoplásicas | ((OG=(“Instituto Nacional de Enfermedades Neoplásicas” OR “National Institute Of Neoplastic Diseases” OR INEN)) **OR** (OO=(“Instituto Nacional de Enfermedades Neoplásicas” OR “National Institute Of Neoplastic Diseases” OR INEN)) **OR** (OO=(“Instituto Nacional de Enfermedades Neoplásicas” OR “National Institute Of Neoplastic Diseases” OR INEN))) NOT ((OG=(INEN NEAR/5 cuba) OR OO=(INEN NEAR/5 cuba) OR AD=(INEN NEAR/5 cuba)) OR (OG=(INEN NEAR/5 endocrinologia) OR OO=(INEN NEAR/5 endocrinologia) OR AD=(INEN NEAR/5 endocrinologia))) |
| #3 | Instituto Nacional de Ciencias Neurológicas | OG=(“Instituto Nacional de Ciencias Neurológicas” OR “National Institute Of Neurological Sciences”) OR OO=(“Instituto Nacional de Ciencias Neurológicas” OR “National Institute Of Neurological Sciences”) OR AD=(“Instituto Nacional de Ciencias Neurológicas” OR “National Institute Of Neurological Sciences”) OR ((OG=(INCN NEAR/5 peru) OR OO=(INCN NEAR/5 peru) OR AD=(INCN NEAR/5 peru)) OR (OG=(INCN NEAR/5 lima) OR OO=(INCN NEAR/5 lima) OR AD=(INCN NEAR/5 lima))) |
| #4 | Instituto Nacional de Oftalmología | ((OG=(“Instituto Nacional de Oftalmología” OR “National Institute Of Ophthalmology”)) **OR** (OO=(“Instituto Nacional de Oftalmología” OR “National Institute Of Ophthalmology”)) **OR** (AD=(“Instituto Nacional de Oftalmología” OR “National Institute Of Ophthalmology”))) OR ((OG=(INO NEAR/5 peru) OR OO=(INO NEAR/5 peru) OR AD=(INO NEAR/5 peru)) OR (OG=(INO NEAR/5 lima) OR OO=(INO NEAR/5 lima) OR AD=(INO NEAR/5 lima))) |
| #5 | Instituto Nacional de Rehabilitación | ((OG=(“Instituto Nacional de Rehabilitación Dra. Adriana Rebaza Flores” OR “National Institute of Rehabilitation Dra. Adriana Rebaza Flores”)) **OR** (OO=(“Instituto Nacional de Rehabilitación Dra. Adriana Rebaza Flores” OR “National Institute of Rehabilitation Dra. Adriana Rebaza Flores”)) **OR** (AD=(“Instituto Nacional de Rehabilitación Dra. Adriana Rebaza Flores” OR “National Institute of Rehabilitation Dra. Adriana Rebaza Flores”))) OR (OG=(“Instituto Nacional de Rehabilitación” NEAR/5 Lima) OR OO=(“Instituto Nacional de Rehabilitación” NEAR/5 Lima) OR AD=(“Instituto Nacional de Rehabilitación” |

| | | |
|---|---|---|
| | | NEAR/5 Lima)) OR (OG=("Instituto Nacional de Rehabilitación" NEAR/5 peru) OR OO=("Instituto Nacional de Rehabilitación" NEAR/5 peru) OR AD=("Instituto Nacional de Rehabilitación" NEAR/5 peru)) OR (OG=(INR NEAR/5 Lima) OR OO=(INR NEAR/5 Lima) OR AD=(INR NEAR/5 Lima)) OR (OG=(INR NEAR/5 peru) OR OO=(INR NEAR/5 peru) OR AD=(INR NEAR/5 peru)) |
| #6 | Instituto Nacional de Salud del Niño de San Borja | (OG=("Instituto Nacional de Salud del Niño - San Borja" OR "Instituto Nacional de Salud del Nino - San Borja" OR "Instituto Nacional de Salud Nino San Borja" OR INSN-SB)) **OR** (OO=("Instituto Nacional de Salud del Nino - San Borja" OR "Instituto Nacional de Salud Nino San Borja" OR INSN-SB)) **OR** (AD=("Instituto Nacional de Salud del Nino - San Borja" OR "Instituto Nacional de Salud Nino San Borja" OR INSN-SB)) |
| #7 | Instituto Nacional de Salud del Niño de Breña | (OG=("Instituto Nacional de Salud del Niño de Breña" OR "Instituto Nacional de Salud Nino de Breña" OR INSNB OR (niño AND breña))) **OR** (OO=("Instituto Nacional de Salud del Niño de Breña" OR "Instituto Nacional de Salud Nino de Breña" OR INSNB OR (niño AND breña))) **OR** (AD=("Instituto Nacional de Salud del Niño de Breña" OR "Instituto Nacional de Salud Nino de Breña" OR INSNB OR (niño AND breña))) |
| #8 | Instituto Nacional Materno Perinatal | ((OG=("Instituto Nacional Materno Perinatal" OR "National Maternal Perinatal Institute")) **OR** (OO=("Instituto Nacional Materno Perinatal" OR "National Maternal Perinatal Institute")) **OR** (AD=("Instituto Nacional Materno Perinatal" OR "National Maternal Perinatal Institute"))) OR (OG=("Instituto Nacional Materno Perinatal" NEAR/5 Lima) OR OO=("Instituto Nacional Materno Perinatal" NEAR/5 Lima) OR AD=("Instituto Nacional Materno Perinatal" NEAR/5 Lima)) OR (OG=("Instituto Nacional Materno Perinatal" NEAR/5 peru) OR OO=("Instituto Nacional Materno Perinatal" NEAR/5 peru) OR AD=("Instituto Nacional Materno Perinatal" NEAR/5 peru)) OR (OG=(INMP NEAR/5 Lima) OR OO=(INMP NEAR/5 Lima) OR AD=(INMP NEAR/5 Lima)) OR (OG=(INMP NEAR/5 peru) OR OO=(INMP NEAR/5 peru) OR AD=(INMP NEAR/5 peru)) |
| #9 | Instituto Nacional de Salud Mental | ((OG=("Instituto Nacional de Salud Mental" OR "Instituto Nacional de Salud Mental Honorio Delgado Hideyo Noguchi" OR "National Institute Of Mental Health" OR "National Institute Of Mental Health Honorio Delgado Hideyo Noguchi")) OR (OO=("Instituto Nacional de Salud Mental" OR "Instituto Nacional de Salud Mental Honorio Delgado Hideyo Noguchi" OR "National Institute Of Mental Health" OR "National Institute Of Mental Health Honorio Delgado Hideyo Noguchi")) OR (AD=("Instituto Nacional de Salud Mental" OR "Instituto Nacional de Salud Mental Honorio Delgado Hideyo Noguchi" OR "National Institute Of Mental Health" OR "National Institute Of Mental Health Honorio Delgado Hideyo Noguchi"))) OR (OG=("Instituto Nacional de Salud Mental" NEAR/5 Lima) OR OO=("Instituto Nacional de Salud Mental" NEAR/5 Lima) OR AD=("Instituto Nacional de Salud Mental" NEAR/5 Lima)) OR (OG=("Instituto Nacional de Salud Mental" NEAR/5 peru) OR OO=("Instituto Nacional de Salud Mental" NEAR/5 peru) OR AD=("Instituto Nacional de Salud Mental" NEAR/5 peru)) OR (OG=(INSM NEAR/5 Lima) OR OO=(INSM NEAR/5 Lima) OR AD=(INSM NEAR/5 Lima)) OR (OG=(INSM NEAR/5 peru) OR OO=(INSM NEAR/5 peru) OR AD=(INSM NEAR/5 peru)) |
| SCIELO SEARCH | | |

| | | |
|---|---|---|
| #1 | Instituto Nacional de Salud | (OG=(INS Peru)) OR OG=(Instituto Nacional de Salud - Peru) OR (OO=(INS Peru)) OR OO=(Instituto Nacional de Salud - Peru) OR (AD=(INS Peru)) OR AD=(Instituto Nacional de Salud - Peru) |
| #2 | Instituto Nacional de Enfermedades Neoplásicas | ((OG=("Instituto Nacional de Enfermedades Neoplásicas" OR "National Institute Of Neoplastic Diseases" OR INEN)) OR (OO=("Instituto Nacional de Enfermedades Neoplásicas" OR "National Institute Of Neoplastic Diseases" OR INEN)) OR (OO=("Instituto Nacional de Enfermedades Neoplásicas" OR "National Institute Of Neoplastic Diseases" OR INEN))) NOT ((OG=(INEN NEAR/5 cuba) OR OO=(INEN NEAR/5 cuba) OR AD=(INEN NEAR/5 cuba)) OR (OG=(INEN NEAR/5 endocrinologia) OR OO=(INEN NEAR/5 endocrinologia) OR AD=(INEN NEAR/5 endocrinologia))) |
| #3 | Instituto Nacional de Ciencias Neurológicas | OG=("Instituto Nacional de Ciencias Neurológicas" OR "National Institute Of Neurological Sciences") OR OO=("Instituto Nacional de Ciencias Neurológicas" OR "National Institute Of Neurological Sciences") OR AD=("Instituto Nacional de Ciencias Neurológicas" OR "National Institute Of Neurological Sciences") OR ((OG=(INCN NEAR/5 peru) OR OO=(INCN NEAR/5 peru) OR AD=(INCN NEAR/5 peru)) OR (OG=(INCN NEAR/5 lima) OR OO=(INCN NEAR/5 lima) OR AD=(INCN NEAR/5 lima))) |
| #4 | Instituto Nacional de Oftalmología | (OG=("Instituto Nacional de Oftalmología" OR "National Institute Of Ophthalmology"OR INO)) **OR** (OO=("Instituto Nacional de Oftalmología" OR "National Institute Of Ophthalmology"OR INO)) **OR** (AD=("Instituto Nacional de Oftalmología" OR "National Institute Of Ophthalmology"OR INO)) |
| #5 | Instituto Nacional de Rehabilitación | ((OG=("Instituto Nacional de Rehabilitación Dra. Adriana Rebaza Flores" OR "National Institute of Rehabilitation Dra. Adriana Rebaza Flores")) **OR** (OO=("Instituto Nacional de Rehabilitación Dra. Adriana Rebaza Flores" OR "National Institute of Rehabilitation Dra. Adriana Rebaza Flores")) **OR** (AD=("Instituto Nacional de Rehabilitación Dra. Adriana Rebaza Flores" OR "National Institute of Rehabilitation Dra. Adriana Rebaza Flores"))) OR (OG=("Instituto Nacional de Rehabilitación" NEAR/5 Lima) OR OO=("Instituto Nacional de Rehabilitación" NEAR/5 Lima) OR AD=("Instituto Nacional de Rehabilitación" NEAR/5 Lima)) OR (OG=("Instituto Nacional de Rehabilitación" NEAR/5 peru) OR OO=("Instituto Nacional de Rehabilitación" NEAR/5 peru) OR AD=("Instituto Nacional de Rehabilitación" NEAR/5 peru)) OR (OG=(INR NEAR/5 Lima) OR OO=(INR NEAR/5 Lima) OR AD=(INR NEAR/5 Lima)) OR (OG=(INR NEAR/5 peru) OR OO=(INR NEAR/5 peru) OR AD=(INR NEAR/5 peru)) |
| #6 | Instituto Nacional de Salud del Niño de San Borja | (OG=("Instituto Nacional de Salud del Niño - San Borja" OR "Instituto Nacional de Salud del Nino - San Borja" OR "Instituto Nacional de Salud Nino San Borja" OR INSN-SB)) OR (OO=("Instituto Nacional de Salud del Nino - San Borja" OR "Instituto Nacional de Salud Nino San Borja" OR INSN-SB)) OR (AD=("Instituto Nacional de Salud del Nino - San Borja" OR "Instituto Nacional de Salud Nino San Borja" OR INSN-SB)) |
| #7 | Instituto Nacional de Salud del Niño de Breña | (OG=("Instituto Nacional de Salud del Niño de Breña" OR "Instituto Nacional de Salud Nino de Breña" OR INSNB OR (niño AND breña))) OR (OO=("Instituto Nacional de Salud del Niño de Breña" OR "Instituto Nacional de Salud Nino de Breña" OR INSNB OR (niño AND breña))) OR (AD=("Instituto Nacional de Salud del Niño de Breña" OR "Instituto Nacional de Salud |

| | | |
|---|---|---|
| | | Nino de Breña” OR INSNB OR (niño AND breña))) |
| #8 | Instituto Nacional Materno Perinatal | ((OG=(“Instituto Nacional Materno Perinatal” OR “National Maternal Perinatal Institute”)) **OR** (OO=(“Instituto Nacional Materno Perinatal” OR “National Maternal Perinatal Institute”)) **OR** (AD=(“Instituto Nacional Materno Perinatal” OR “National Maternal Perinatal Institute”))) OR (OG=(“Instituto Nacional Materno Perinatal” NEAR/5 Lima) OR OO=(“Instituto Nacional Materno Perinatal” NEAR/5 Lima) OR AD=(“Instituto Nacional Materno Perinatal” NEAR/5 Lima)) OR (OG=(“Instituto Nacional Materno Perinatal” NEAR/5 peru) OR OO=(“Instituto Nacional Materno Perinatal” NEAR/5 peru) OR AD=(“Instituto Nacional Materno Perinatal” NEAR/5 peru)) OR (OG=(INMP NEAR/5 Lima) OR OO=(INMP NEAR/5 Lima) OR AD=(INMP NEAR/5 Lima)) OR (OG=(INMP NEAR/5 peru) OR OO=(INMP NEAR/5 peru) OR AD=(INMP NEAR/5 peru)) |
| #9 | Instituto Nacional de Salud Mental | (OG=(“Instituto Nacional de Salud Mental” OR “Instituto Nacional de Salud Mental Honorio Delgado Hideyo Noguchi” OR “National Institute Of Mental Health” OR “National Institute Of Mental Health Honorio Delgado Hideyo Noguchi” OR INSM)) **OR** (OO=(“Instituto Nacional de Salud Mental” OR “Instituto Nacional de Salud Mental Honorio Delgado Hideyo Noguchi” OR “National Institute Of Mental Health” OR “National Institute Of Mental Health Honorio Delgado Hideyo Noguchi” OR INSM)) **OR** (AD=(“Instituto Nacional de Salud Mental” OR “Instituto Nacional de Salud Mental Honorio Delgado Hideyo Noguchi” OR “National Institute Of Mental Health” OR “National Institute Of Mental Health Honorio Delgado Hideyo Noguchi” OR INSM)) |
| EMBASE SEARCH | | |
| #1 | Instituto Nacional de Salud 60071247 | (('Instituto Nacional De Salud' OR 'Instituto Nacional De Salud. Lima' OR 'Instituto Nacional De Salud. Lima' OR 'Instituto Nacional De Salud (ins)' OR 'Instituto National De Salud' OR 'National Institute Of Health' OR 'Instituto Nacional De Salud Del Peru') NEAR/5 (Peru OR Lima)):ff OR ('instituto nacional de salud, lima' OR 'instituto nacional de salud, peru'):ff OR (('ins' NEAR/5 ('lima' OR 'peru')):ff) |
| #2 | Instituto Nacional de Enfermedades Neoplásicas 60071241 | (('Instituto Nacional De Enfermedades Neoplásicas' OR 'Instituto Nacional De Enfermedades Neoplasicas' OR 'Instituto Nacional De Enfermedades Neoplásicas (inen)' OR 'National Institute Of Neoplastic Diseases' OR 'Instituto Nacional De Enfermedades Neoplásicas. Lima' OR 'Instituto De Enfermedades Neoplásicas' OR 'Instituto De Enfermedades Neoplasicas' OR 'Instituto Nacional De Enfermedades Neoplasicas (inen)' OR 'National Cancer Institute Of Peru (inen)' OR 'inen') NEAR/5 (Peru OR Lima)):ff OR ('Instituto Nacional de Enfermedades Neoplasicas' OR 'Instituto Nacional de Enfermedades Neoplasicas, lima' OR 'Instituto Nacional de Enfermedades Neoplasicas, peru'):ff OR (('inen' NEAR/5 ('lima' OR 'peru')):ff) |
| #3 | Instituto Nacional de Ciencias Neurológicas | (('Instituto Nacional De Ciencias Neurológicas' OR 'Instituto Nacional De Ciencias Neurologicas' OR 'Instituto De Ciencias Neurologicas' OR 'Instituto De Ciencias Neurológicas' OR 'National Institute Of Neurological Sciences' OR 'Inst. Nac. De Ciencias Neurologicas' OR 'Inst. Especializado En Cie. Neurol.' OR 'Instituto Especializado En Ciencias Neurológicas' OR 'Inst. Nac. De Cie. Neurologicas' OR 'Inst. De Cie. Neurológicas') NEAR/5 (Peru OR Lima)):ff **OR** ('Instituto Nacional De Ciencias Neurológicas' OR 'Instituto Nacional De Ciencias Neurológicas, lima' OR 'Instituto Nacional De Ciencias Neurológicas, peru'):ff |

| | | |
|---|---|---|
| | | **OR** (('incn' NEAR/5 ('lima' OR 'peru')):ff) |
| #4 | Instituto Nacional de Oftalmología | (('Instituto Nacional De Oftalmología' OR 'Instituto Nacional De Oftalmología Dr. Francisco Contreras Campos' OR 'Instituto Nacional De Oftalmologia' OR 'National Institute Of Ophthalmology' OR 'Instituto De Oftalmología (ino)' OR 'Instituto De Oftalmologia (ino)' OR 'Ocular Pathology Laboratory dr. José Antonio Avendaño Valdez' OR 'Instituto Nacional De Oftalmología dr. Francisco Contreras Campos' OR 'National Institute Of Ophthalmology Of Peru' OR 'Instituto Nacional De Oftalmología francisco Contreras Campos' OR 'Instituto Nacional De Oftalmología francisco Contreras Campos') NEAR/5 (Peru OR Lima)):ff OR ('Instituto Nacional De Oftalmología' OR 'Instituto Nacional De Oftalmología, lima' OR 'Instituto Nacional De Oftalmología, peru'):ff OR (('ino' NEAR/5 ('lima' OR 'peru')):ff) |
| #5 | Instituto Nacional de Rehabilitación | (('Instituto Nacional de Rehabilitación' OR 'Instituto Nacional de Rehabilitación Dra. Adriana Rebaza Flores' OR 'National Institute of Rehabilitation' OR 'National Institute of Rehabilitation Dra. Adriana Rebaza Flores') NEAR/5 (Peru OR Lima)):ff OR (('Instituto Nacional de Rehabilitación' NEAR/5 ('lima' OR 'peru')) OR 'Instituto Nacional de Rehabilitación, lima' OR 'Instituto Nacional de Rehabilitación, peru'):ff OR ((inr NEAR/5 ('lima' OR 'peru')):ff) |
| #6 | Instituto Nacional de Salud del Niño de San Borja | (('Instituto Nacional de Salud del Niño de San Borja' OR 'Instituto Nacional De Salud Del Niño-san Borja' OR 'Instituto Nacional De Salud Del Niño De San Borja' OR 'Instituto Nacional De Salud Del Niño San Borja (insn-sb)' OR 'Instituto Nacional De Salud Del Niño — San Borja' OR 'Instituto Nacional De Salud Del Niño - San Borja' OR 'Instituto Nacional De Salud Del Nino San Borja' OR 'Cuidados Paliativos Del Instituto Nacional De Salud Del Niño De San Borja' OR 'Instituto Nacional Del Niño San Borja' OR 'Instituto Nacional Del Niño') NEAR/5 (Peru OR Lima)):ff OR (('instituto nacional de salud del niño de san borja' OR ('niño' AND 'san borja')) OR 'Instituto Nacional de Salud del Niño de San Borja, lima' OR 'Instituto Nacional de Salud del Niño de San Borja, peru'):ff OR ((('INSN-SB' OR 'INSNSB') NEAR/5 ('lima' OR 'peru')):ff) |
| #7 | Instituto Nacional de Salud del Niño de Breña | (('Instituto Nacional De Salud Del Niño' OR 'Instituto De Salud Del Niño' OR 'National Institute Of Child Health' OR 'Instituto Nacional De Salud Del Niño-breña' OR 'Instituto Nacional De Salud Del Nino, Lima' OR 'Instituto Nacional De Salud Del Nino' OR 'Instituto Nacional De Salud Del Niño. Lima' OR 'Instituto Nacional De Salud Del Niño Breña' OR 'Instituto De Salud Del Nino Hosp. Nino') NEAR/5 (Peru OR Lima)):ff OR (('instituto nacional de salud del niño de breña' OR ('niño' AND 'breña')) OR 'Instituto Nacional de Salud del Niño de Breña, lima' OR 'Instituto Nacional de Salud del Niño de Breña, peru'):ff OR ((('INSN-Breña' OR 'INSNB' ) NEAR/5 ('lima' OR 'peru')):ff) |
| #7 | Instituto Nacional de Salud del Niño de Breña | (('Instituto Nacional De Salud Del Niño breña' OR 'Instituto Nacional De Salud Del Nino breña, Lima' OR 'Instituto Nacional De Salud Del Niño breña. Lima' OR 'Instituto Nacional De Salud Del Niño Breña' OR 'Instituto De Salud Del Nino Hosp. Nino breña') NEAR/5 (Peru OR Lima)):ff OR (('instituto nacional de salud del niño de breña' OR ('niño' AND 'breña')) OR 'Instituto Nacional de Salud del Niño de Breña, lima' OR 'Instituto Nacional de Salud del Niño de Breña, peru'):ff OR ((('INSN-Breña' OR 'INSNB' ) NEAR/5 ('lima' OR 'peru')):ff) |
| #8 | Instituto Nacional Materno Perinatal | (('Instituto Nacional Materno Perinatal' OR 'Instituto Nacional Materno Perinatal De Lima' OR 'Instituto Nacional Materno-perinatal' OR 'Instituto Nacional Materno Perinatal Inmp' OR 'Instituto Nacional Materno Perinatal' OR 'Servicio De Medicina |

| | | |
|---|---|---|
| | | Fetal Del Instituto Nacional Materno Perinatal' OR 'Servicio De Cuidados Intensivos Del Instituto Nacional Materno Perinatal' OR 'National Maternal Perinatal Institute' OR 'National Institute For Maternal Perinatal Health (inmp)' OR 'Instituto Nacional Materno Perinatal. Universidad Privada Del Norte. Lima') NEAR/5 (Peru OR Lima)):ff OR ('Instituto Nacional Materno Perinatal' OR 'Instituto Nacional Materno Perinatal, lima' OR 'Instituto Nacional Materno Perinatal, peru'):ff OR (('INMP' NEAR/5 ('lima' OR 'peru')):ff) |
| #9 | Instituto Nacional de Salud Mental | (('Instituto Nacional De Salud Mental Honorio Delgado-hideyo Noguchi' OR 'National Institute Of Mental Health Honorio Delgado Hideyo Noguchi' OR 'Peruvian National Institute Of Mental Health' OR 'National Institute Of Mental Health' OR 'Instituto Nacional De Salud Mental honorio Delgado - Hideyo Noguchi') NEAR/5 (Peru OR Lima)):ff OR ('Instituto Nacional de Salud Mental' OR 'Instituto Nacional de Salud Mental, lima' OR 'Instituto Nacional de Salud Mental, peru' OR 'Instituto Nacional de Salud Mental Honorio Delgado-Hideyo Noguchi' OR 'Instituto Nacional de Salud Mental Honorio Delgado-Hideyo Noguchi, lima' OR 'Instituto Nacional de Salud Mental Honorio Delgado-Hideyo Noguchi, peru'):ff OR (('INSM' NEAR/5 ('lima' OR 'peru')):ff) |

**Supplementary Material 2.** Funding of original articles from specialized institutes in Peru.

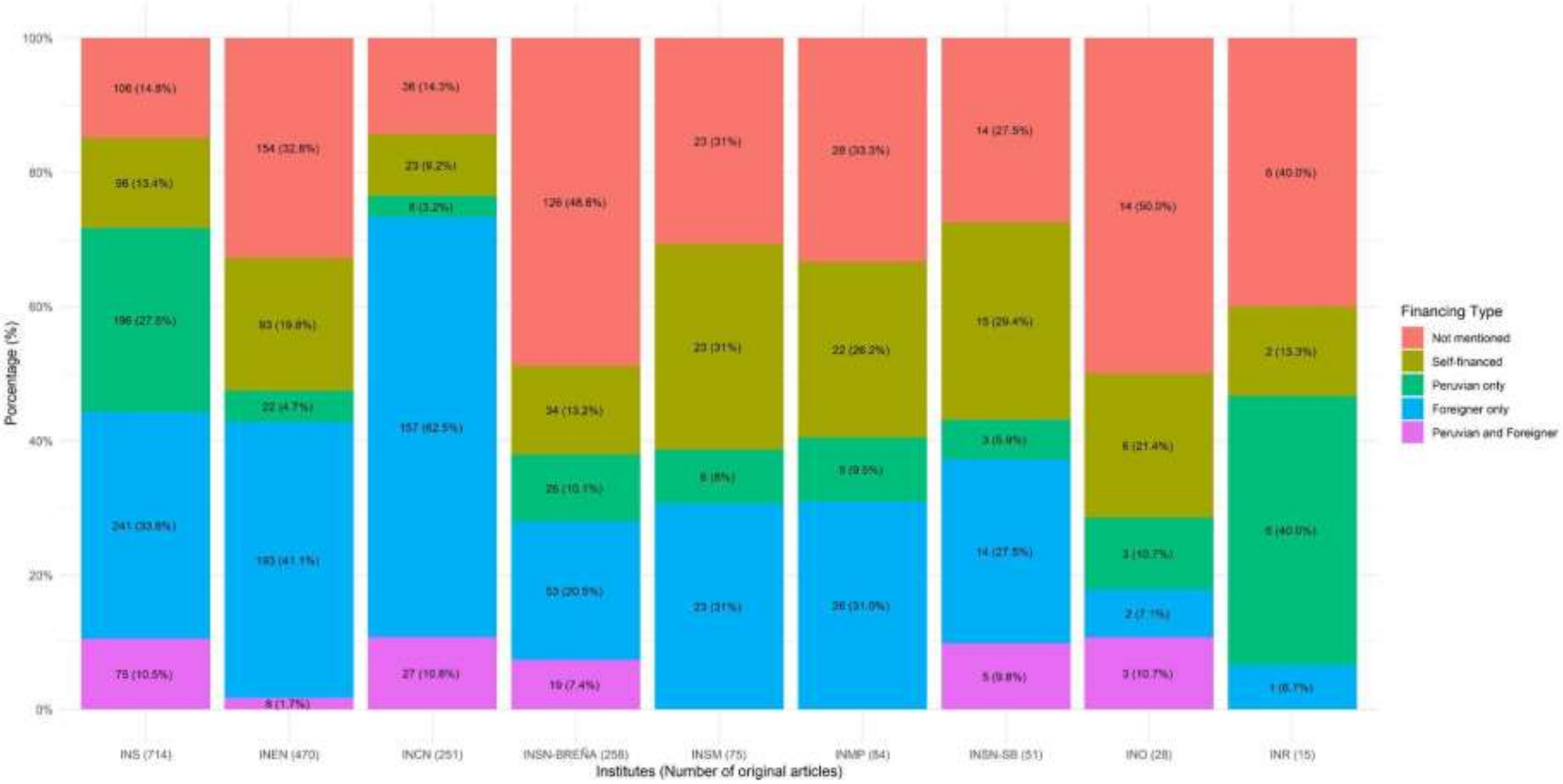

**Supplementary Material 3.** Journal characteristics and specialization by specialized health institutes in Peru.

| Institute | Top 3 journals | | | | |
|---|---|---|---|---|---|
| | **Top** | **Name of journal** | **Percentage of production n / N (%)** | **Quartil*** | **Country*** |
| **Global** | 1 | Revista Peruana de Medicina Experimental y Salud Publica | 510 / 3200 (16,89) | Q3 | Peru |
| | 2 | Revista de Neuro-Psiquiatria | 93 / 3200 (3,08) | No quartile | Peru |
| | 3 | Revista de Gastroenterologia del Peru | 89 / 3200 (2,95) | Q4 | Peru |
| **INS** | 1 | Revista Peruana de Medicina Experimental y Salud Pública | 354 / 1049 (33,75) | Q3 | Peru |
| | 2 | The Lancet | 42 / 1049 (4) | Q1 | Others |
| | 3 | Anales de la Facultad de Medicina | 34 / 1049 (3,24) | No quartile | Peru |
| **INEN** | 1 | Revista de Gastroenterologia del Peru | 65 / 722 (9) | Q4 | Peru |
| | 2 | Revista Peruana de Medicina Experimental y Salud Publica | 38 / 722 (5,26) | Q3 | Peru |
| | 3 | Journal of Clinical Oncology | 25 / 722 (3,46) | Q1 | Others |
| **INCN** | 1 | American Journal of Tropical Medicine and Hygiene | 38 / 413 (9,2) | Q1 | Others |
| | 2 | Revista de Neuro-Psiquiatria | 29 / 413 (7,02) | No quartile | Peru |
| | 3 | PLoS Neglected Tropical Diseases | 28 / 413 (6,78) | Q1 | Others |
| **INSN-BREÑA** | 1 | Revista Peruana de Medicina Experimental y Salud Pública | 62 / 390 (15,9) | Q3 | Peru |
| | 2 | Revista de Gastroenterologia del Peru | 11 / 390 (2,82) | Q4 | Peru |

| | | | | | |
|---|---|---|---|---|---|
| | 3 | American Journal of Tropical Medicine and Hygiene | 6 / 390 (1,54) | Q1 | Others |
| **INSM** | 1 | Revista de Neuro-Psiquiatría | 52 / 153 (33,99) | No quartile | Peru |
| | 2 | Revista Peruana de Medicina Experimental y Salud Publica | 15 / 153 (9,8) | Q3 | Peru |
| | 3 | Acta Medica Peruana | 8 / 153 (5,23) | No quartile | Peru |
| **INMP** | 1 | Revista Peruana de Ginecologia y Obstetricia | 12 / 113 (10,62) | No quartile | Peru |
| | 2 | Revista Peruana de Medicina Experimental y Salud Publica | 9 / 113 (7,96) | Q3 | Peru |
| | 3 | Colombian Journal of Anestesiology | 4 / 113 (3,54) | Q3 | Others |
| **INSN-SB** | 1 | Revista Peruana de Medicina Experimental y Salud Pública | 12 / 106 (11,32) | Q3 | Peru |
| | 2 | Pediatric Radiology | 5 / 106 (4,72) | Q2 | Otros |
| | 3 | Actas Dermo-Sifiliograficas | 4 / 106 (3,77) | Q3 | Otros |
| **INO** | 1 | Archivos de la Sociedad Espanola de Oftalmologia | 5 / 40 (12,5) | Q4 | Otros |
| | 2 | Revista Mexicana de Oftalmología | 4 / 40 (10) | Q4 | Latin America |
| | 3 | Revista Peruana de Medicina Experimental y Salud Pública | 4 / 40 (10) | Q3 | Peru |
| **INR** | 1 | Revista Medica Herediana | 22 / 34 (64,71) | No quartile | Peru |
| | 2 | Revista de Neuro-Psiquiatría | 3 / 34 (8,82) | No quartile | Peru |
| | 3 | Anales de la Facultad de Medicina | 2 / 34 (5,88) | No quartile | Peru |

**Supplementary Material 4.** Analysis of most researched specialties globally and by institute.

| **Institute** | **Top 1 Specialty** | |
|---|---|---|
| | **Specialty** | **Percent of production**<br>**n / N (%)** |
| **Global** | Medicina de enfermedades infecciosas y tropicales | 937 / 3020 (31,03) |
| **INS** | Medicina de enfermedades infecciosas y tropicales | 522 / 1049 (49,76) |
| **INEN** | Oncología | 620 / 722 (85,87) |
| **INCN** | Neurología | 334 / 413 (80,87) |
| **INSN-BREÑA** | Pediatría | 172 / 390 (44,1) |
| **INSM** | Psiquiatría | 136 / 153 (88,89) |
| **INMP** | Ginecología y obstetricia | 68 / 113 (60,18) |
| **INSN-SB** | Medicina de enfermedades infecciosas y tropicales | 35 / 106 (33,02) |
| **INO** | Oftalmología | 37 / 40 (92,5) |
| **INR** | Medicina física y de rehabilitación | 27 / 34 (79,41) |

**Supplementary Material 5.** Global and specialized institute word maps.

Global

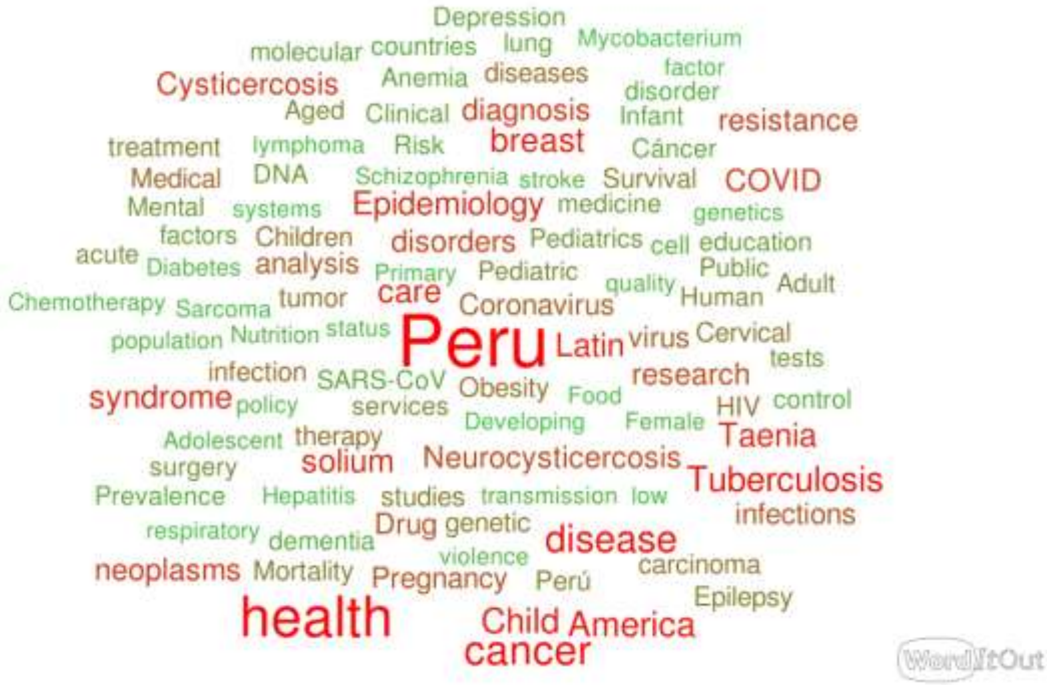

INS

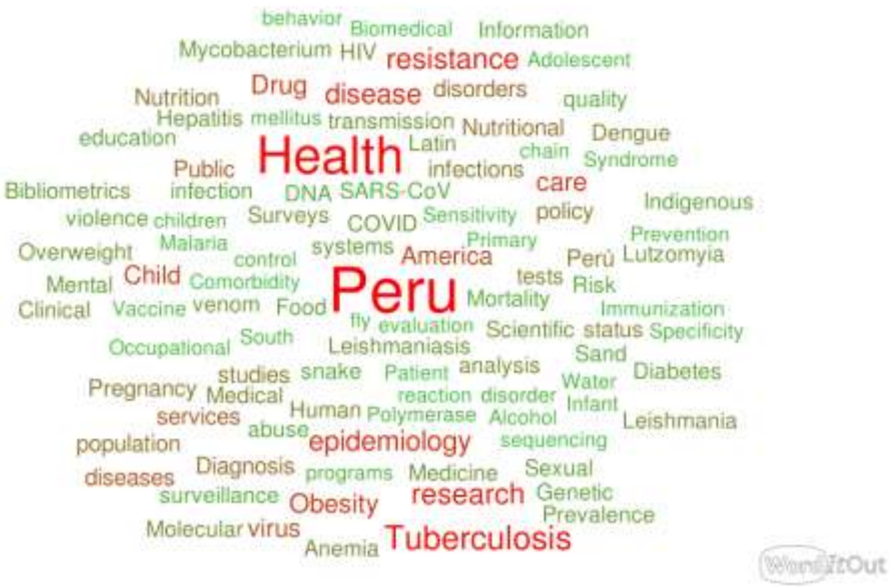

behavior Biomedical Information
Mycobacterium HIV resistance Adolescent
Nutrition Drug disease disorders quality
Hepatitis mellitus transmission Nutritional
education Health Latin Dengue
Public infections chain Syndrome
Bibliometrics infection DNA SARS-CoV care Indigenous
violence children Surveys COVID Sensitivity policy
Overweight Malaria control systems America Primary Prevention Perú Lutzomyia
Mental Child Comorbidity Peru tests Risk
Clinical Vaccine venom Food Mortality Immunization
fly evaluation Scientific status Specificity
Occupational South Leishmaniasis Sand
studies snake Patient analysis Diabetes
Pregnancy Medical reaction disorder Water
services Human Polymerase Alcohol Infant Leishmania
population abuse epidemiology sequencing
diseases Diagnosis programs Medicine Sexual
surveillance Obesity research Genetic
Molecular virus Prevalence
Anemia Tuberculosis
WordItOut

INEN

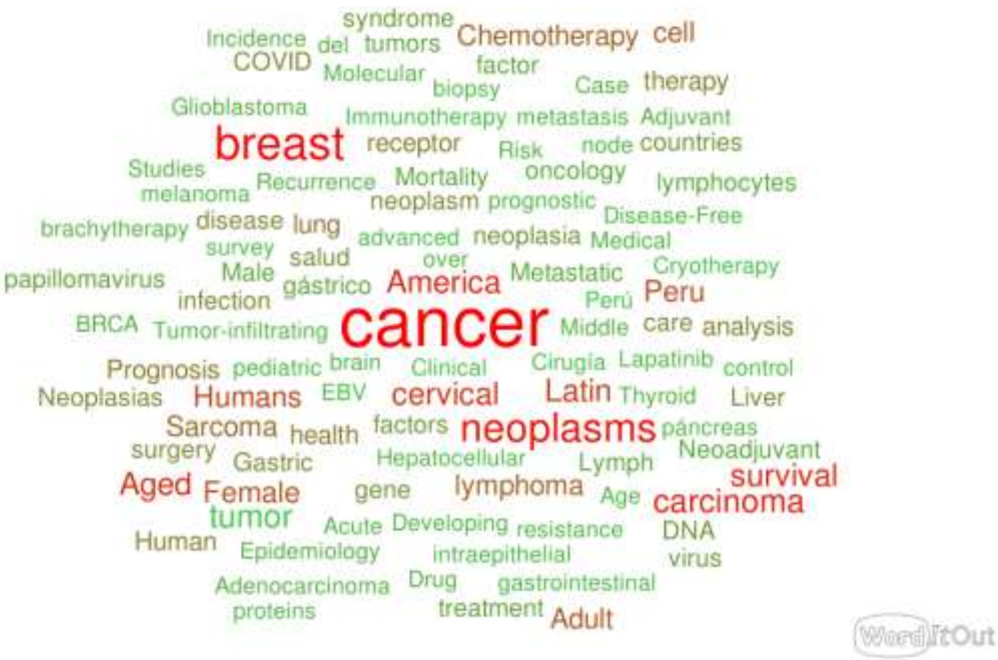

syndrome
Incidence del tumors Chemotherapy cell
COVID Molecular factor
biopsy Case therapy
Glioblastoma Immunotherapy metastasis Adjuvant
breast receptor Risk node countries
Studies Mortality oncology lymphocytes
melanoma Recurrence neoplasm prognostic
brachytherapy disease lung Disease-Free
survey advanced neoplasia Medical
salud over
papillomavirus Male gástrico America Metastatic Cryotherapy
infection Perú Peru
BRCA Tumor-infiltrating cancer Middle care analysis
Prognosis pediatric brain Clinical Cirugia Lapatinib control
Neoplasias Humans EBV cervical Latin Thyroid Liver
Sarcoma health factors neoplasms páncreas
surgery Gastric Hepatocellular Lymph Neoadjuvant
Aged Female gene lymphoma Age carcinoma survival
tumor Acute Developing resistance DNA
Human Epidemiology intraepithelial virus
Adenocarcinoma Drug gastrointestinal
proteins treatment Adult
WordItOut

INCN

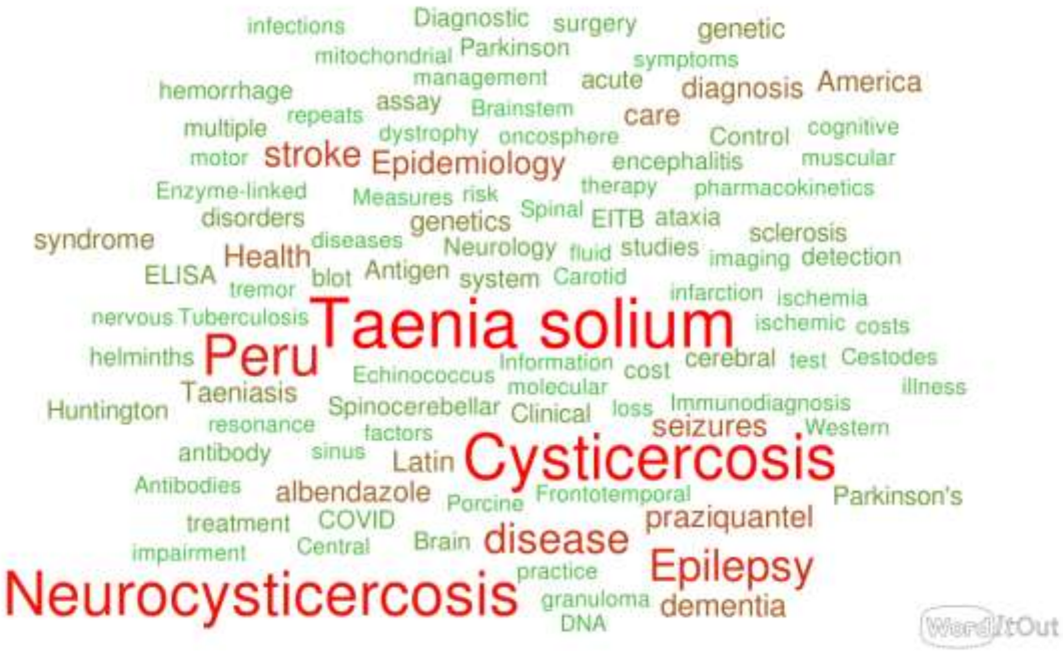
infections
Diagnostic
surgery
genetic
mitochondrial
Parkinson
symptoms
hemorrhage
management
acute
diagnosis
America
repeats
assay
Brainstem
care
multiple
dystrophy
oncosphere
Control
cognitive
motor
stroke
Epidemiology
encephalitis
muscular
Enzyme-linked
Measures
risk
therapy
pharmacokinetics
disorders
genetics
Spinal
EITB
ataxia
syndrome
diseases
Neurology
fluid
studies
sclerosis
Health
imaging
detection
ELISA
tremor
blot
Antigen
system
Carotid
infarction
ischemia
nervous
Tuberculosis
Taenia solium
ischemic
costs
helminths
Peru
Information
cost
cerebral
test
Cestodes
Echinococcus
molecular
illness
Taeniasis
Huntington
Spinocerebellar
Clinical
loss
Immunodiagnosis
resonance
factors
seizures
Western
antibody
sinus
Latin
Cysticercosis
Antibodies
albendazole
Porcine
Frontotemporal
Parkinson's
treatment
COVID
praziquantel
impairment
Central
Brain
disease
Epilepsy
practice
Neurocysticercosis
granuloma
dementia
DNA
WordItOut

INSN-Breña

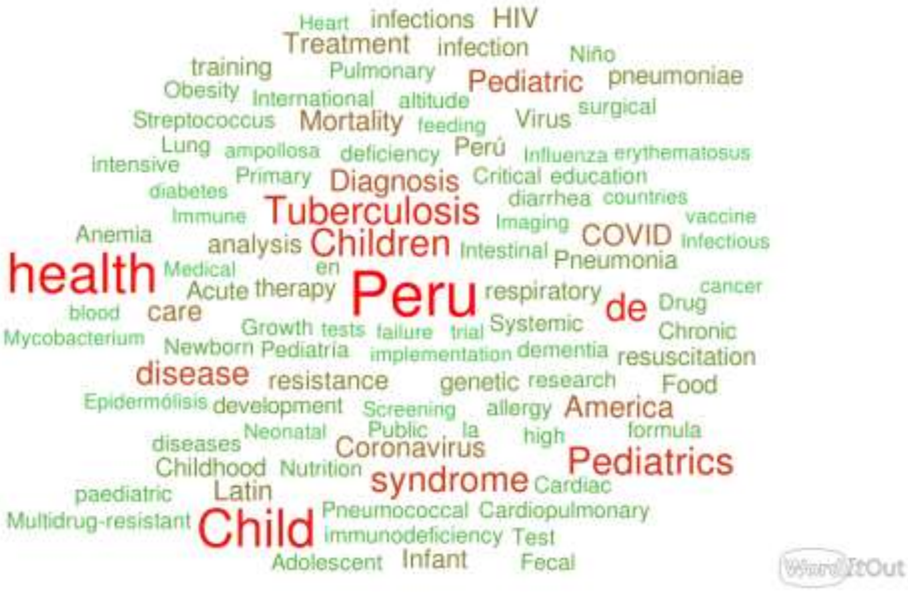
Heart infections HIV
Treatment infection
training Niño
Pulmonary Pediatric pneumoniae
Obesity International altitude
Streptococcus Mortality feeding Virus surgical
Lung ampollosa deficiency Perú Influenza erythematosus
intensive
Primary Diagnosis Critical education
diabetes diarrhea countries
Immune Tuberculosis Imaging vaccine
Anemia COVID
analysis Children Intestinal Pneumonia Infectious
health Medical en
Acute therapy Peru respiratory cancer
de Drug
blood care
Growth tests failure trial Systemic Chronic
Mycobacterium
Newborn Pediatría implementation dementia resuscitation
disease resistance genetic research Food
Epidermólisis development Screening allergy America
Neonatal Public la high formula
diseases Coronavirus
Childhood Nutrition syndrome Cardiac Pediatrics
paediatric Latin
Pneumococcal Cardiopulmonary
Multidrug-resistant Child immunodeficiency Test
Adolescent Infant Fecal
WordItOut

INSM

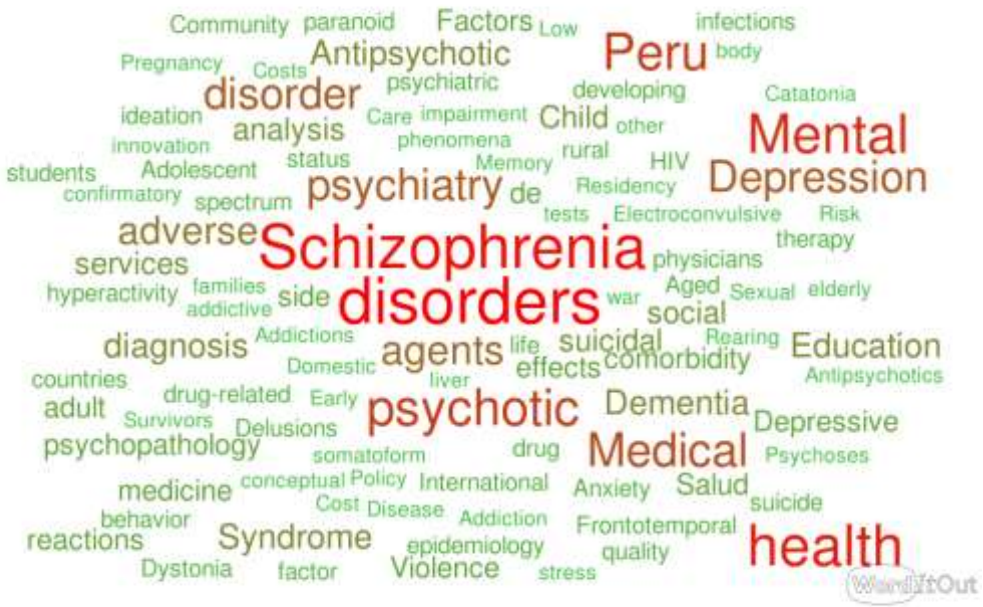
Community paranoid Factors Low infections
Pregnancy Costs Antipsychotic Peru body
disorder psychiatric developing Catatonia
ideation Care impairment Child other Mental
innovation analysis phenomena
students Adolescent status Memory rural HIV Depression
confirmatory spectrum psychiatry de Residency
tests Electroconvulsive Risk
adverse Schizophrenia therapy
services physicians
hyperactivity families addictive side disorders war Aged Sexual elderly social
diagnosis Addictions agents life suicidal Rearing Education
Domestic effects comorbidity
countries liver Antipsychotics
drug-related Early Dementia
adult Survivors Delusions psychotic Depressive
psychopathology somatoform drug Medical Psychoses
medicine conceptual Policy International Anxiety Salud
Cost Disease Addiction suicide
behavior Frontotemporal
reactions Syndrome epidemiology quality health
Dystonia factor Violence stress
WordItOut

INMP

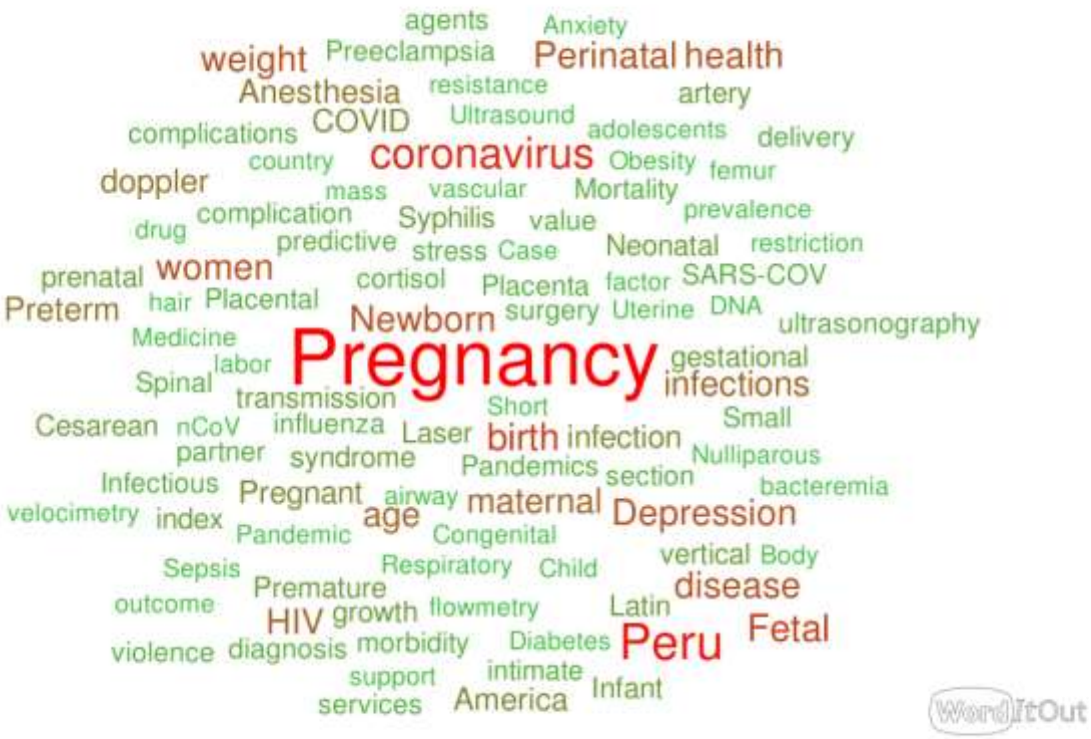
agents
Anxiety
weight
Preeclampsia
Perinatal
health
Anesthesia
resistance
artery
complications
COVID
Ultrasound
adolescents
delivery
country
coronavirus
Obesity
femur
doppler
mass
vascular
Mortality
drug
complication
Syphilis
value
prevalence
predictive
stress
Case
Neonatal
restriction
prenatal
women
cortisol
Placenta
factor
SARS-COV
Preterm
hair
Placental
Newborn
surgery
Uterine
DNA
ultrasonography
Medicine
Pregnancy
gestational
labor
Spinal
infections
transmission
Short
Cesarean
nCoV
influenza
Laser
birth
infection
Small
partner
syndrome
Pandemics
Nulliparous
section
Infectious
Pregnant
airway
maternal
bacteremia
velocimetry
index
age
Depression
Pandemic
Congenital
vertical
Body
Sepsis
Respiratory
Child
Premature
disease
outcome
HIV
growth
flowmetry
Latin
Fetal
violence
diagnosis
morbidity
Diabetes
Peru
support
intimate
services
America
Infant
WordItOut

INSN-SB

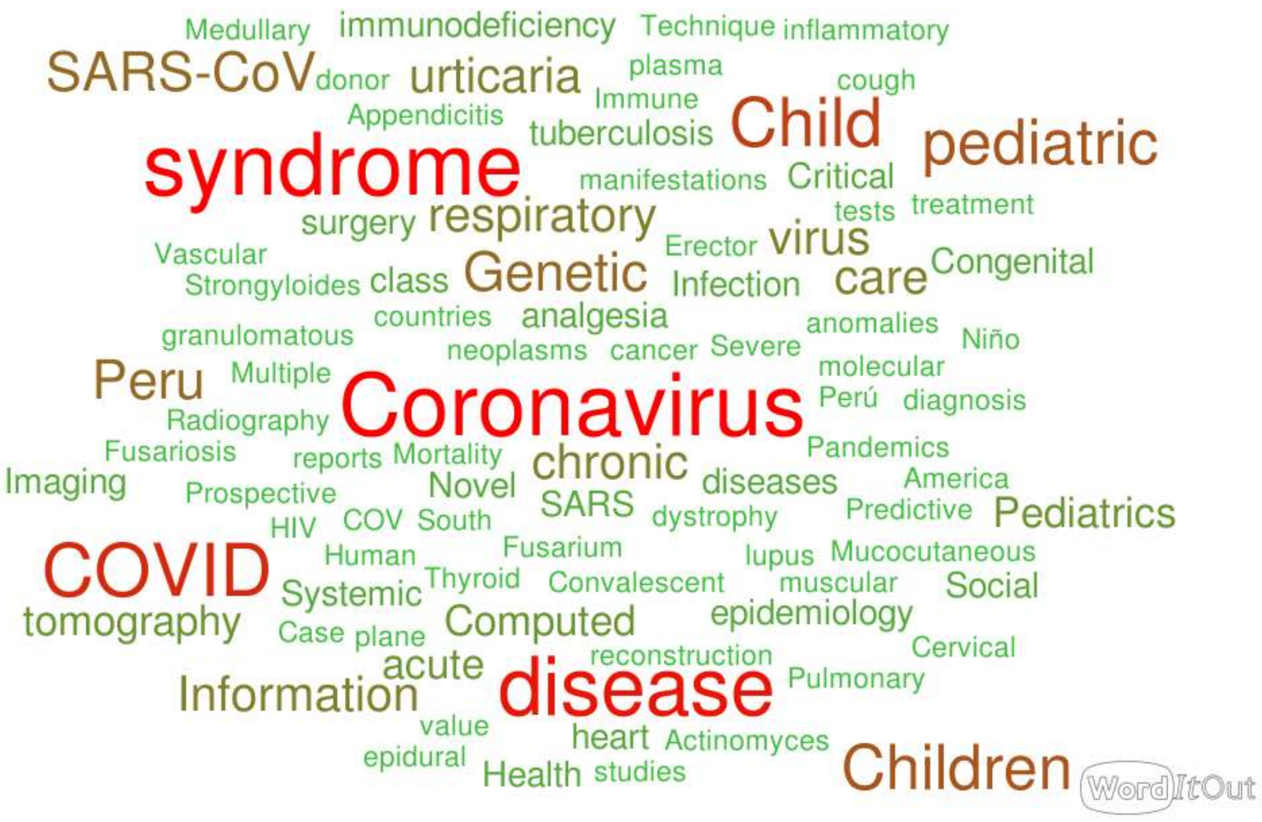

Medullary
immunodeficiency
Technique
inflammatory
SARS-CoV
donor
urticaria
plasma
cough
Immune
Appendicitis
tuberculosis
Child
pediatric
syndrome
manifestations
Critical
surgery
respiratory
tests
treatment
Erector
virus
Vascular
Congenital
Strongyloides
class
Genetic
Infection
care
countries
analgesia
anomalies
granulomatous
neoplasms
cancer
Severe
Niño
Peru
Multiple
molecular
Coronavirus
Perú
diagnosis
Radiography
Fusariosis
Pandemics
reports
Mortality
chronic
Imaging
diseases
America
Prospective
Novel
SARS
HIV
COV
South
dystrophy
Predictive
Pediatrics
Fusarium
Human
lupus
Mucocutaneous
COVID
Thyroid
Convalescent
muscular
Social
Systemic
tomography
Computed
epidemiology
Case
plane
reconstruction
Cervical
acute
Pulmonary
Information
disease
value
heart
Actinomyces
epidural
Health
studies
Children
WordItOut

INO

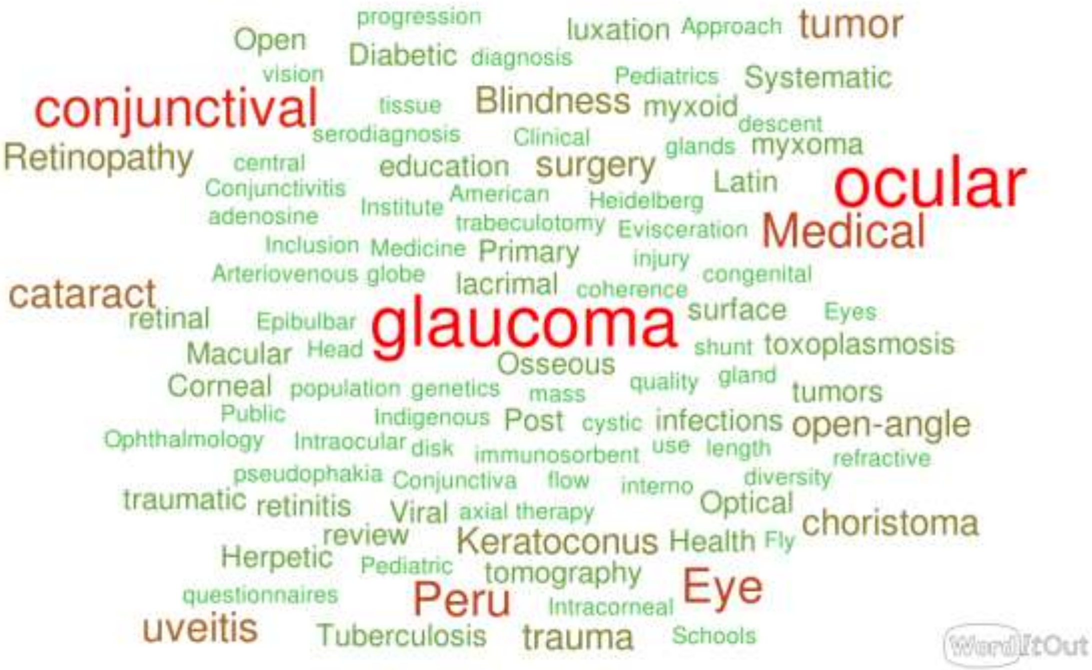

progression
Open
luxation
Approach
tumor
Diabetic
diagnosis
vision
Pediatrics
Systematic
conjunctival
tissue
Blindness
myxoid
serodiagnosis
Clinical
descent
glands
myxoma
Retinopathy
central
education
surgery
Conjunctivitis
American
Latin
ocular
adenosine
Institute
Heidelberg
trabeculotomy
Evisceration
Medical
Inclusion
Medicine
Primary
injury
Arteriovenous
globe
lacrimal
coherence
congenital
cataract
retinal
Epibulbar
glaucoma
surface
Eyes
Macular
Head
Osseous
shunt
toxoplasmosis
Corneal
population
genetics
mass
quality
gland
tumors
Public
Indigenous
Post
cystic
infections
open-angle
Ophthalmology
Intraocular
disk
immunosorbent
use
length
refractive
pseudophakia
Conjunctiva
flow
interno
diversity
traumatic
retinitis
Viral
axial
therapy
Optical
choristoma
review
Keratoconus
Health
Fly
Herpetic
Pediatric
tomography
Eye
questionnaires
Peru
Intracorneal
uveitis
Tuberculosis
trauma
Schools
WordItOut

INR

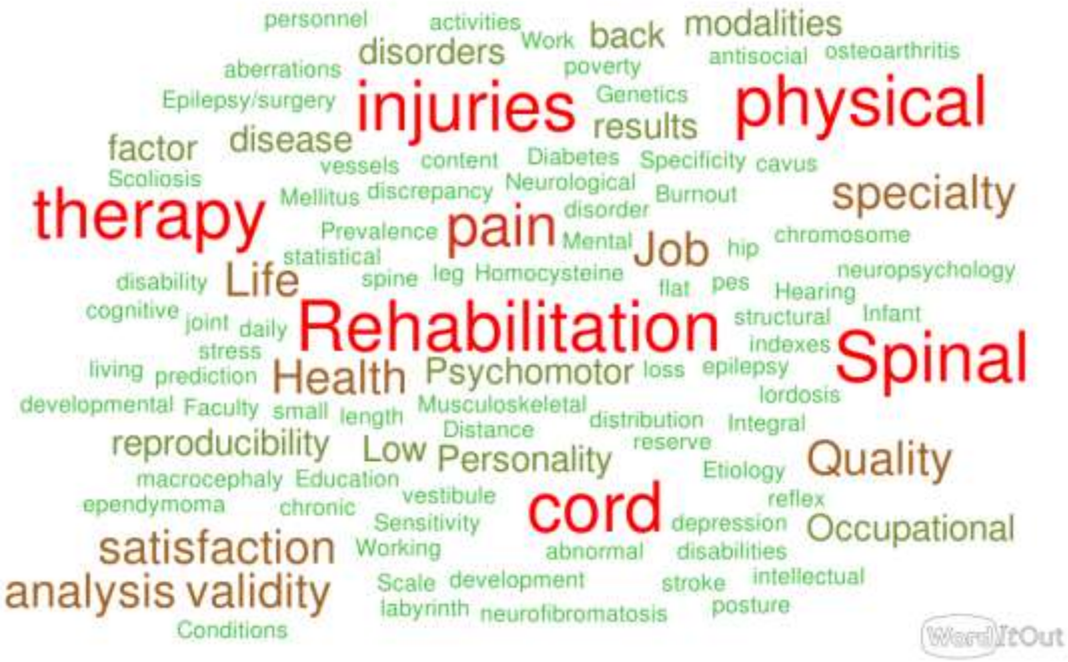
personnel
activities
Work
back
modalities
disorders
antisocial
osteoarthritis
aberrations
poverty
Epilepsy/surgery
injuries
Genetics
physical
results
factor
disease
vessels
content
Diabetes
Specificity
cavus
Scoliosis
Mellitus
discrepancy
Neurological
Burnout
specialty
therapy
disorder
Prevalence
pain
Mental
Job
hip
chromosome
statistical
disability
Life
spine
leg
Homocysteine
flat
pes
neuropsychology
Hearing
cognitive
joint
daily
Rehabilitation
structural
Infant
stress
indexes
Spinal
living
prediction
Health
Psychomotor
loss
epilepsy
lordosis
developmental
Faculty
small
length
Musculoskeletal
distribution
Integral
Distance
reproducibility
Low
Personality
reserve
Quality
Etiology
macrocephaly
Education
vestibule
reflex
ependymoma
chronic
Sensitivity
cord
depression
Occupational
satisfaction
Working
abnormal
disabilities
Scale
development
stroke
intellectual
analysis
validity
labyrinth
neurofibromatosis
posture
Conditions
WordItOut

**Supplementary Material 6.** Number of publications by specialized institutes in Peru in subperiods from 1991-2021.

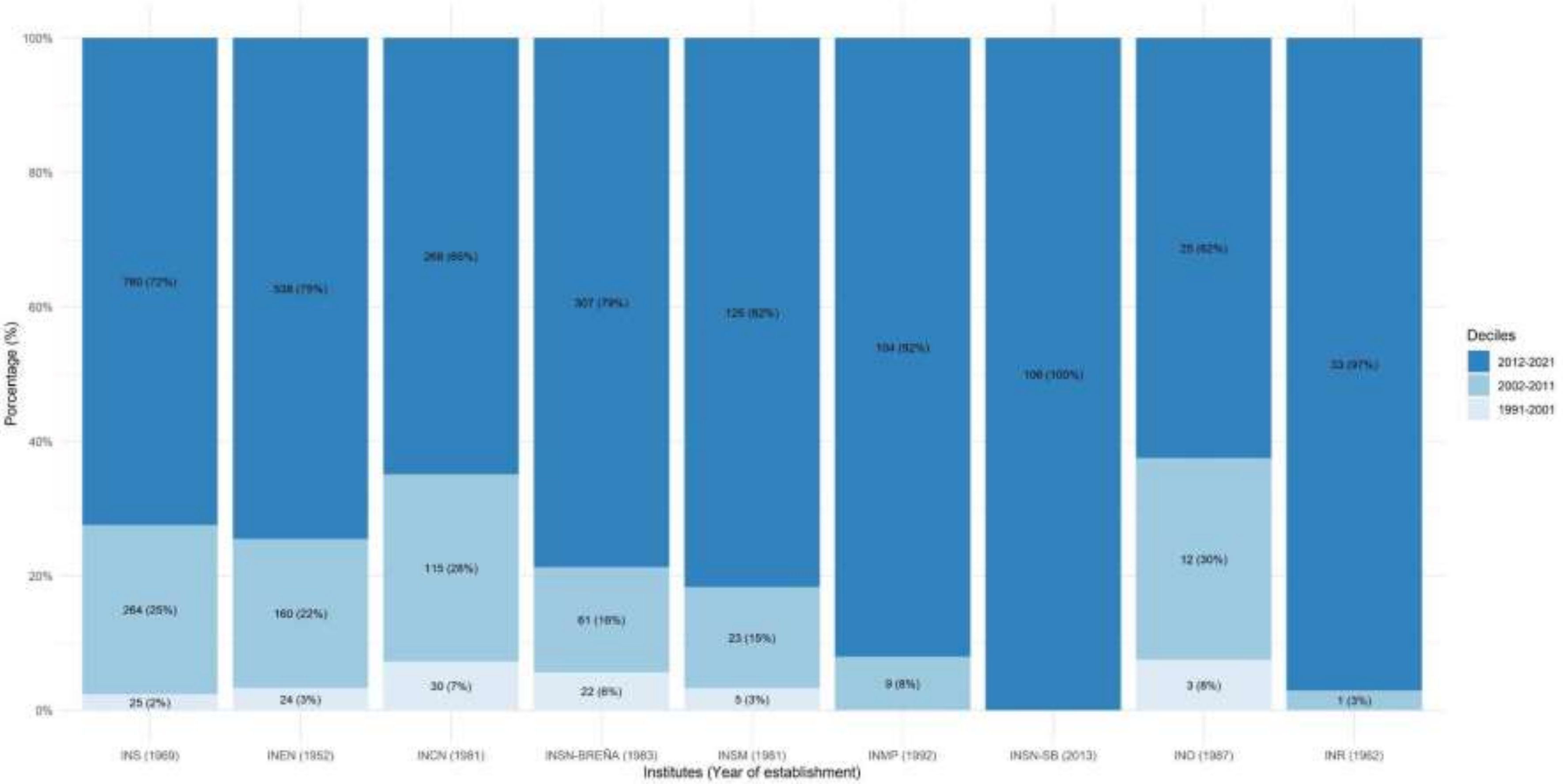

**Supplementary Material 7.** Production indicators for each institute.

| **Institute** | Total production in 2020 | Total production in 2021 | Productivity variation rate (2020 - 2021) | Total publications by author by institute | Percentage of production per author by institute | Percentage of publications by author according to institute of total publications | Productivity index by Institute | Productivity index by gender (of the first authorship of the institutes) Leadership by first authorship | Productivity index by gender (of the last authorship of the institutes) Leadership by last authorship |
|---|---|---|---|---|---|---|---|---|---|
| **INS** | 78 | 81 | 3,85 | 183 | 17,45 | 17.44 | 6,96 | 0,56 | 0,35 |
| **INEN** | 96 | 96 | 0 | 245 | 33,93 | 33.93 | 6,58 | 0,31 | 0,18 |
| **INCN** | 31 | 54 | 74,19 | 319 | 77,24 | 77.24 | 6,02 | 0,16 | 0,16 |
| **INSN-Breña** | 51 | 65 | 27,45 | 88 | 22,56 | 22.56 | 5,97 | 0,29 | 0,47 |
| **INSM** | 25 | 16 | -36 | 3 | 1,96 | 1.96 | 5,03 | 0,4 | 0,14 |
| **INMP** | 29 | 21 | -27,59 | 52 | 46,02 | 18.87 | 4,73 | 0,53 | 0,41 |
| **INSN-SB** | 25 | 39 | 56 | 20 | 18,87 | 18.87 | 4,66 | 0,67 | 0,48 |
| **INO** | 6 | 4 | -33,33 | 15 | 37,5 | 37.5 | 3,69 | 0,64 | 0,88 |
| **INR** | 3 | 1 | -66,67 | 15 | 44,12 | 44.18 | 3,53 | 1,31 | 1,57 |

**Supplementary Material 8.** Collaboration diagram among specialized institutes in Peru.

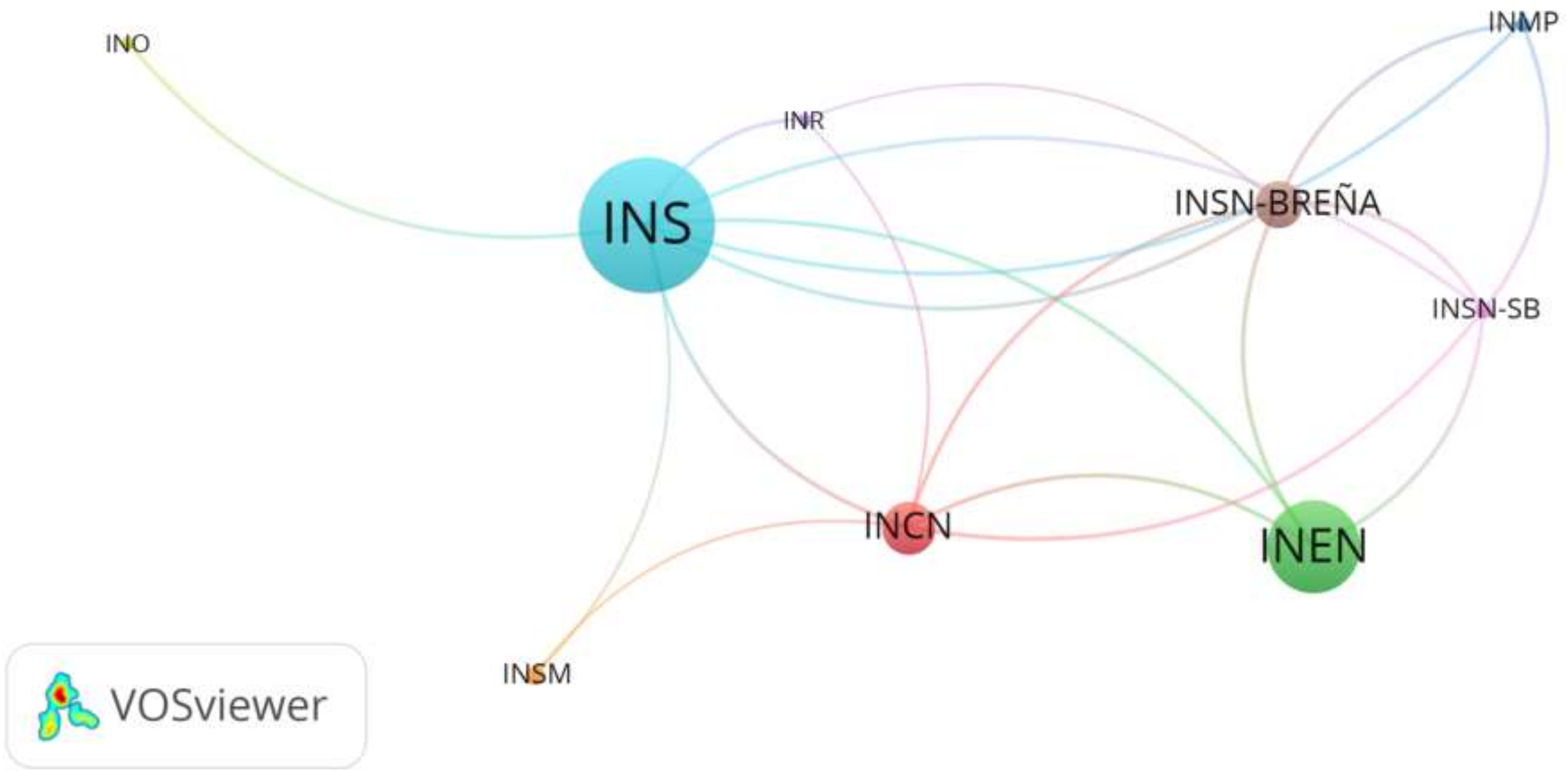

**Supplementary Material 8.** Stratified gender analysis of first and last authorship.

| **Institute** | **First authorship** | | **Gender of first authorship** | | **No first authorship** | | **With first authorship** | | **Last authorship** | | **Gender of last authorship** | | **No last authorship** | | **With last authorship** | |
|---|---|---|---|---|---|---|---|---|---|---|---|---|---|---|---|---|
| | **No N=1712** | **Yes N=1308** | **Female N=921** | **Male N=2099** | **Female N=542** | **Male N=1170** | **Female N=379** | **Male N=929** | **No N=1748** | **Si N=1176** | **Female N=677** | **Male N=2247** | **Female N=408** | **Male N=1340** | **Female N=269** | **Male N=907** |
| **INS** | 597 (56.9%) | 452 (43.1%) | 339 (32.3%) | 710 (67.7%) | 177 (29.6%) | 420 (70.4%) | 162 (35.8%) | 290 (64.2%) | 597 (58.5%) | 424 (41.5%) | 230 (22.5%) | 791 (77.5%) | 120 (29.4%) | 477 (35.6%) | 110 (40.9%) | 314 (34.6%) |
| **INEN** | 422 (58.4%) | 300 (41.6%) | 197 (27.3%) | 525 (72.7%) | 126 (29.9%) | 296 (70.1%) | 71 (23.7%) | 229 (76.3%) | 427 (60.5%) | 279 (39.5%) | 131 (18.6%) | 575 (81.4%) | 89 (21.8%) | 338 (25.2%) | 42 (15.6%) | 237 (26.1%) |
| **INCN** | 254 (61.5%) | 159 (38.5%) | 90 (21.8%) | 323 (78.2%) | 68 (26.8%) | 186 (73.2%) | 22 (13.8%) | 137 (86.2%) | 260 (63.0%) | 153 (37.0%) | 69 (16.7%) | 344 (83.3%) | 48 (11.8%) | 212 (15.8%) | 21 (7.81%) | 132 (14.6%) |
| **INSN-Breña** | 240 (61.5%) | 150 (38.5%) | 139 (35.6%) | 251 (64.4%) | 105 (43.8%) | 135 (56.2%) | 34 (22.7%) | 116 (77.3%) | 241 (63.1%) | 141 (36.9%) | 125 (32.7%) | 257 (67.3%) | 80 (19.6%) | 161 (12.0%) | 45 (16.7%) | 96 (10.6%) |
| **INSM** | 55 (35.9%) | 98 (64.1%) | 48 (31.4%) | 105 (68.6%) | 20 (36.4%) | 35 (63.6%) | 28 (28.6%) | 70 (71.4%) | 66 (50.4%) | 65 (49.6%) | 22 (16.8%) | 109 (83.2%) | 14 (3.43%) | 52 (3.88%) | 8 (2.97%) | 57 (6.28%) |
| **INMP** | 67 (59.3%) | 46 (40.7%) | 43 (38.1%) | 70 (61.9%) | 27 (40.3%) | 40 (59.7%) | 16 (34.8%) | 30 (65.2%) | 68 (62.4%) | 41 (37.6%) | 52 (47.7%) | 57 (52.3%) | 40 (9.80%) | 28 (2.09%) | 12 (4.46%) | 29 (3.20%) |
| **INSN-SB** | 51 (48.1%) | 55 (51.9%) | 34 (32.1%) | 72 (67.9%) | 12 (23.5%) | 39 (76.5%) | 22 (40.0%) | 33 (60.0%) | 61 (60.4%) | 40 (39.6%) | 25 (24.8%) | 76 (75.2%) | 12 (2.94%) | 49 (3.66%) | 13 (4.83%) | 27 (2.98%) |
| **INO** | 22 (55.0%) | 18 (45.0%) | 12 (30.0%) | 28 (70.0%) | 5 (22.7%) | 17 (77.3%) | 7 (38.9%) | 11 (61.1%) | 22 (59.5%) | 15 (40.5%) | 12 (32.4%) | 25 (67.6%) | 5 (1.23%) | 17 (1.27%) | 7 (2.60%) | 8 (0.88%) |
| **INR** | 4 (11.8%) | 30 (88.2%) | 19 (55.9%) | 15 (44.1%) | 2 (50.0%) | 2 (50.0%) | 17 (56.7%) | 13 (43.3%) | 6 (25.0%) | 18 (75.0%) | 11 (45.8%) | 13 (54.2%) | 0 (0.00%) | 6 (0.45%) | 11 (4.09%) | 7 (0.77%) |

**Supplementary Material 10.** Association models for highly cited and quartile 1 publications.

**A. Highly cited bivariate analysis**

| | **Normal case** | | | **Minimum case** | | | **Maximum case** | | |
|---|---|---|---|---|---|---|---|---|---|
| | **No N=1470** | **Yes N=1117** | **P-value** | **No N=1903** | **Yes N=1117** | **P-value** | **No N=1470** | **Yes N=1550** | **P-value** |
| **Language:** | | | <0.001 | | | <0.001 | | | <0.001 |
| **Spanish** | 589 (86.87%) | 89 (13.13%) | | 920 (91.18%) | 89 (8.82%) | | 589 (58.37%) | 420 (41.63%) | |
| **Spanish and English** | 124 (91.18%) | 12 (8.82%) | | 141 (92.16%) | 12 (7.84%) | | 124 (81.05%) | 29 (18.95%) | |
| **English** | 755 (42.66%) | 1015 (57.34%) | | 839 (45.25%) | 1015 (54.75%) | | 755 (40.72%) | 1099 (59.28%) | |
| **Others** | 2 (66.67%) | 1 (33.33%) | | 3 (75.00%) | 1 (25.00%) | | 2 (50.00%) | 2 (50.00%) | |
| **Study Type** | | | <0.001 | | | <0.001 | | | <0.001 |
| **Basic** | 121 (41.72%) | 169 (58.28%) | | 143 (45.83%) | 169 (54.17%) | | 121 (38.78%) | 191 (61.22%) | |
| **Interventional** | 43 (26.88%) | 117 (73.12%) | | 57 (32.76%) | 117 (67.24%) | | 43 (24.71%) | 131 (75.29%) | |
| **Not applicable** | 601 (78.87%) | 161 (21.13%) | | 795 (83.16%) | 161 (16.84%) | | 601 (62.87%) | 355 (37.13%) | |
| **Observational** | 579 (51.74%) | 540 (48.26%) | | 737 (57.71%) | 540 (42.29%) | | 579 (45.34%) | 698 (54.66%) | |
| **Secondary** | 126 | 130 | | 171 | 130 | | 126 | 175 | |

| | | | | | | | | | |
|---|---|---|---|---|---|---|---|---|---|
| | (49.22%) | (50.78%) | | (56.81%) | (43.19%) | | (41.86%) | (58.14%) | |
| **Country of the journal:** | | | <0.001 | | | <0.001 | | | <0.001 |
| **Peru** | 516 (87.02%) | 77 (12.98%) | | 851 (91.70%) | 77 (8.30%) | | 516 (55.60%) | 412 (44.40%) | |
| **Latin America** | 179 (84.83%) | 32 (15.17%) | | 199 (86.15%) | 32 (13.85%) | | 179 (77.49%) | 52 (22.51%) | |
| **Others** | 775 (43.47%) | 1008 (56.53%) | | 853 (45.84%) | 1008 (54.16%) | | 775 (41.64%) | 1086 (58.36%) | |
| **Cross-cutting theme:** | | | <0.001 | | | <0.001 | | | 0.072 |
| **No** | 1392 (56.08%) | 1090 (43.92%) | | 1792 (62.18%) | 1090 (37.82%) | | 1392 (48.30%) | 1490 (51.70%) | |
| **Yes** | 78 (74.29%) | 27 (25.71%) | | 111 (80.43%) | 27 (19.57%) | | 78 (56.52%) | 60 (43.48%) | |
| **Funding:** | | | <0.001 | | | <0.001 | | | <0.001 |
| **No** | 927 (70.02%) | 397 (29.98%) | | 1280 (76.33%) | 397 (23.67%) | | 927 (55.28%) | 750 (44.72%) | |
| **Yes** | 543 (42.99%) | 720 (57.01%) | | 623 (46.39%) | 720 (53.61%) | | 543 (40.43%) | 800 (59.57%) | |
| **Original article:** | | | <0.001 | | | <0.001 | | | <0.001 |
| **No** | 642 (74.83%) | 216 (25.17%) | | 858 (79.89%) | 216 (20.11%) | | 642 (59.78%) | 432 (40.22%) | |
| **Yes** | 828 | 901 | | 1045 | 901 | | 828 | 1118 | |

| | | | | | | | | | |
|---|---|---|---|---|---|---|---|---|---|
| | (47.89%) | (52.11%) | | (53.70%) | (46.30%) | | (42.55%) | (57.45%) | |
| **Collaboration:** | | | <0.001 | | | <0.001 | | | <0.001 |
| **Individual** | 421 (84.03%) | 80 (15.97%) | | 587 (88.01%) | 80 (11.99%) | | 421 (63.12%) | 246 (36.88%) | |
| **Peruvian** | 542 (79.71%) | 138 (20.29%) | | 751 (84.48%) | 138 (15.52%) | | 542 (60.97%) | 347 (39.03%) | |
| **Foreign** | 507 (36.06%) | 899 (63.94%) | | 565 (38.59%) | 899 (61.41%) | | 507 (34.63%) | 957 (65.37%) | |
| **Quartile of the journal:** | | | <0.001 | | | <0.001 | | | <0.001 |
| **No quartile** | 30 (78.95%) | 8 (21.05%) | | 339 (97.69%) | 8 (2.31%) | | 30 (8.65%) | 317 (91.35%) | |
| **Q4** | 258 (94.51%) | 15 (5.49%) | | 273 (94.79%) | 15 (5.21%) | | 258 (89.58%) | 30 (10.42%) | |
| **Q3** | 640 (82.69%) | 134 (17.31%) | | 692 (83.78%) | 134 (16.22%) | | 640 (77.48%) | 186 (22.52%) | |
| **Q2** | 202 (53.87%) | 173 (46.13%) | | 216 (55.53%) | 173 (44.47%) | | 202 (51.93%) | 187 (48.07%) | |
| **Q1** | 340 (30.17%) | 787 (69.83%) | | 383 (32.74%) | 787 (67.26%) | | 340 (29.06%) | 830 (70.94%) | |
| **First authorship of the institute:** | | | <0.001 | | | <0.001 | | | <0.001 |
| **No** | 690 (44.43%) | 863 (55.57%) | | 849 (49.59%) | 863 (50.41%) | | 690 (40.30%) | 1022 (59.70%) | |

| | | | | | | | | | |
|---|---|---|---|---|---|---|---|---|---|
| **Yes** | 780 (75.44%) | 254 (24.56%) | | 1054 (80.58%) | 254 (19.42%) | | 780 (59.63%) | 528 (40.37%) | |
| **Gender of the first authorship:** | | | 0.103 | | | 0.058 | | | 0.297 |
| **Female** | 462 (59.31%) | 317 (40.69%) | | 604 (65.58%) | 317 (34.42%) | | 462 (50.16%) | 459 (49.84%) | |
| **Male** | 1008 (55.75%) | 800 (44.25%) | | 1299 (61.89%) | 800 (38.11%) | | 1008 (48.02%) | 1091 (51.98%) | |
| **Last authorship of the institute:** | | | <0.001 | | | <0.001 | | | <0.001 |
| **No** | 695 (44.10%) | 881 (55.90%) | | 867 (49.60%) | 881 (50.40%) | | 695 (39.76%) | 1053 (60.24%) | |
| **Yes** | 732 (76.09%) | 230 (23.91%) | | 946 (80.44%) | 230 (19.56%) | | 732 (62.24%) | 444 (37.76%) | |
| **Gender of the last authorship:** | | | <0.001 | | | <0.001 | | | 0.003 |
| **Female** | 365 (64.15%) | 204 (35.85%) | | 473 (69.87%) | 204 (30.13%) | | 365 (53.91%) | 312 (46.09%) | |
| **Male** | 1062 (53.94%) | 907 (46.06%) | | 1340 (59.64%) | 907 (40.36%) | | 1062 (47.26%) | 1185 (52.74%) | |

**B. Highly cited with Poisson regression analysis with robust variance**

| | **Normal case** | | | **Minimum case** | | | **Maximum case** | | |
|---|---|---|---|---|---|---|---|---|---|
| | **PR** | **CI (95%)** | **P-value** | **PR** | **CI (95%)** | **P-value** | **PR** | **CI (95%)** | **P-value** |

| | | | | | | | | | |
|---|---|---|---|---|---|---|---|---|---|
| **Language:** | | | | | | | | | |
| **Spanish** | ref | | | | | | | | |
| **Spanish and English** | 0.52 | 0.29 - 0.95 | **0.032** | 0.560 | 0.31 - 1.02 | 0.060 | 0.642 | 0.48 - 0.87 | **0.004** |
| **English** | 1.27 | 0.9 - 1.8 | 0.177 | 1.368 | 0.97 - 1.93 | 0.077 | 1.034 | 0.83 - 1.29 | 0.768 |
| **Others** | 1.19 | 0.26 - 5.44 | 0.821 | 1.303 | 0.28 - 6.07 | 0.736 | 1.191 | 0.51 - 2.78 | 0.687 |
| **Study Type:** | | | | | | | | | |
| **Basic** | | | | | | | | | |
| **Interventional** | 1.12 | 0.99 - 1.26 | 0.067 | 1.088 | 0.96 - 1.24 | 0.195 | 1.093 | 0.98 - 1.22 | 0.114 |
| **Not applicable** | 0.55 | 0.42 - 0.71 | **0.000** | 0.553 | 0.42 - 0.72 | **0.000** | 0.667 | 0.54 - 0.83 | **0.000** |
| **Observational** | 1.01 | 0.92 - 1.11 | 0.827 | 1.017 | 0.92 - 1.13 | 0.747 | 0.983 | 0.9 - 1.08 | 0.715 |
| **Secondary** | 1.03 | 0.87 - 1.2 | 0.755 | 1.054 | 0.89 - 1.24 | 0.532 | 0.939 | 0.81 - 1.08 | 0.391 |
| **Country of the journal:** | | | | | | | | | |
| **Peru** | | | | | | | | | |
| **Latin America** | 0.86 | 0.55 - 1.34 | 0.508 | 0.923 | 0.59 - 1.45 | 0.730 | 0.774 | 0.58 - 1.03 | 0.076 |
| **Others** | 0.98 | 0.65 - 1.49 | 0.926 | 1.054 | 0.69 - 1.62 | 0.810 | 0.881 | 0.67 - 1.15 | 0.349 |

| | | | | | | | | | |
|---|---|---|---|---|---|---|---|---|---|
| **Cross-cutting theme:** | | | | | | | | | |
| **No** | | | | | | | | | |
| **Yes** | 0.77 | 0.56 - 1.05 | 0.099 | 0.789 | 0.57 - 1.09 | 0.145 | 0.845 | 0.7 - 1.01 | 0.071 |
| **Funding:** | | | | | | | | | |
| **No** | | | | | | | | | |
| **Yes** | 1.05 | 0.96 - 1.15 | 0.290 | 1.079 | 0.98 - 1.18 | 0.109 | 0.997 | 0.93 - 1.07 | 0.945 |
| **Original article:** | | | | | | | | | |
| **No** | | | | | | | | | |
| **Yes** | 0.83 | 0.67 - 1.02 | 0.075 | 0.825 | 0.67 - 1.02 | 0.077 | 0.904 | 0.76 - 1.08 | 0.260 |
| **Collaboration:** | | | | | | | | | |
| **Individual** | | | | | | | | | |
| **Peruvian** | 0.99 | 0.77 - 1.28 | 0.957 | 0.984 | 0.76 - 1.27 | 0.900 | 0.997 | 0.87 - 1.14 | 0.968 |
| **Foreign** | 1.42 | 1.1 - 1.84 | **0.007** | 1.452 | 1.12 - 1.88 | **0.004** | 1.328 | 1.13 - 1.57 | **0.001** |
| **Quartile of the journal:** | | | | | | | | | |
| **No quartile** | | | | | | | | | |
| **Q4** | 0.34 | 0.16 - 0.73 | **0.005** | 1.915 | 0.79 - 4.62 | 0.148 | 0.124 | 0.09 - 0.18 | **0.000** |
| **Q3** | 1.00 | 0.56 - 1.8 | 0.993 | 5.604 | 2.72 - 11.53 | **0.000** | 0.247 | 0.21 - 0.29 | **0.000** |
| **Q2** | 1.53 | 0.85 - 2.74 | 0.152 | 8.100 | 3.67 - 17.87 | **0.000** | 0.421 | 0.34 - 0.53 | **0.000** |

| | | | | | | | | | |
|---|---|---|---|---|---|---|---|---|---|
| **Q1** | 2.06 | 1.15 - 3.66 | **0.015** | 10.744 | 4.86 - 23.74 | **0.000** | 0.571 | 0.46 - 0.71 | **0.000** |
| **First authorship of the institute:** | | | | | | | | | |
| **No** | | | | | | | | | |
| **Yes** | 0.88 | 0.78 - 0.98 | **0.025** | 0.869 | 0.77 - 0.98 | 0.018 | 0.960 | 0.88 - 1.04 | 0.339 |
| **Gender of the first authorship:** | | | | | | | | | |
| **Female** | | | | | | | | | |
| **Male** | 1.11 | 1.02 - 1.2 | **0.018** | 1.108 | 1.02 - 1.21 | **0.019** | 1.049 | 0.98 - 1.12 | 0.155 |
| **Last authorship of the institute:** | | | | | | | | | |
| **No** | | | | | | | | | |
| **Yes** | 0.91 | 0.8 - 1.03 | 0.122 | 0.916 | 0.81 - 1.04 | 0.173 | 0.926 | 0.85 - 1.01 | 0.096 |
| **Gender of the last authorship:** | | | | | | | | | |
| **Female** | | | | | | | | | |
| **Male** | 1.11 | 1 - 1.23 | 0.057 | 1.091 | 0.98 - 1.21 | 0.108 | 1.086 | 1 - 1.17 | **0.037** |

### C. Publication in quartile 1 bivariate analysis

| | No N=1850 | Yes N=1170 | P-value |
|---|---|---|---|
| **Language:** | | | 0.000 |
| **Spanish** | 1004 (99.50%) | 5 (0.50%) | |
| **Spanish and English** | 151 (98.69%) | 2 (1.31%) | |
| **English** | 691 (37.27%) | 1163 (62.73%) | |
| **Others** | 4 (100.00%) | 0 (0.00%) | |
| **Study type:** | | | <0.001 |
| **Basic** | 133 (42.63%) | 179 (57.37%) | |
| **Interventional** | 45 (25.86%) | 129 (74.14%) | |
| **Not applicable** | 745 (77.93%) | 211 (22.07%) | |
| **Observational** | 736 (57.64%) | 541 (42.36%) | |
| **Secondary** | 191 (63.46%) | 110 (36.54%) | |
| **Cross-cutting theme:** | | | <0.001 |
| **No** | 1741 (60.41%) | 1141 (39.59%) | |
| **Yes** | 109 (78.99%) | 29 (21.01%) | |
| **Funding:** | | | <0.001 |
| **No** | 1279 (76.27%) | 398 (23.73%) | |

| | | | |
|---|---|---|---|
| **Yes** | 571 (42.52%) | 772 (57.48%) | |
| **Original article:** | | | <0.001 |
| **No** | 821 (76.44%) | 253 (23.56%) | |
| **Yes** | 1029 (52.88%) | 917 (47.12%) | |
| **Collaboration:** | | | <0.001 |
| **Individual** | 594 (89.06%) | 73 (10.94%) | |
| **Peruvian** | 771 (86.73%) | 118 (13.27%) | |
| **Foreign** | 485 (33.13%) | 979 (66.87%) | |
| **First authorship of the institute:** | | | <0.001 |
| **No** | 782 (45.68%) | 930 (54.32%) | |
| **Yes** | 1068 (81.65%) | 240 (18.35%) | |
| **Gender of the first authorship:** | | | 0.450 |
| **Female** | 574 (62.32%) | 347 (37.68%) | |
| **Male** | 1276 (60.79%) | 823 (39.21%) | |
| **Last authorship of the institute:** | | | <0.001 |
| **No** | 802 (45.88%) | 946 (54.12%) | |
| **Yes** | 956 (81.29%) | 220 (18.71%) | |
| **Gender of the last authorship:** | | | <0.001 |

| | | |
|---|---|---|
| **Female** | 471 (69.57%) | 206 (30.43%) |
| **Male** | 1287 (57.28%) | 960 (42.72%) |

**D. Publication in quartile 1 with poisson regression analysis with robust variance**

| | **PR** | **CI (95%)** | **P-value** |
|---|---|---|---|
| **Language:** | | | |
| **Spanish** | ref. | | |
| **Spanish and English** | 2.49 | 0.49 - 12.75 | 0.27 |
| **English** | 71.92 | 29.6 - 174.74 | **0.00** |
| **Others** | 0.00 | 0 - 0 | 0.00 |
| **Study type:** | | | |
| **Basic** | ref. | | |
| **Interventional** | 1.17 | 1.05 - 1.3 | **0.01** |
| **Not applicable** | 0.95 | 0.72 - 1.26 | 0.73 |
| **Observational** | 1.03 | 0.94 - 1.12 | 0.58 |
| **Secondary** | 0.95 | 0.8 - 1.14 | 0.58 |
| **Cross-cutting theme:** | | | |
| **No** | ref. | | |

| | | | |
|---|---|---|---|
| **Yes** | 0.99 | 0.81 - 1.22 | 0.95 |
| **Funding:** | | | |
| **No** | ref. | | |
| **Yes** | 1.23 | 1.13 - 1.34 | **0.00** |
| **Original article:** | | | |
| **No** | ref. | | |
| **Yes** | 1.00 | 0.79 - 1.28 | 0.99 |
| **Collaboration:** | | | |
| **Individual** | ref. | | |
| **Peruvian** | 0.79 | 0.61 - 1.01 | 0.06 |
| **Foreign** | 1.27 | 1.01 - 1.6 | **0.04** |
| **First authorship of the institute:** | | | |
| **No** | ref. | | |
| **Yes** | 0.83 | 0.74 - 0.92 | **0.00** |
| **Gender of the first authorship:** | | | |
| **Female** | ref. | | |
| **Male** | 1.03 | 0.96 - 1.1 | 0.47 |

| | | | |
|---|---|---|---|
| **Last authorship of the institute:** | | | |
| **No** | ref. | | |
| **Yes** | 0.88 | 0.78 - 0.98 | **0.02** |
| **Gender of the last authorship:** | | | |
| **Female** | ref. | | |
| **Male** | 1.16 | 1.06 - 1.28 | **0.00** |
